# Diabetes and the Life-Course: Evidence from Panel Data and Electronic Health Records

**DOI:** 10.64898/2026.06.06.26355069

**Authors:** Chris Heitzig, David Rehkopf

**Author notes:** Corresponding author: Chris Heitzig.

## Abstract

Diabetes screening can alter both clinical care and patient behavior, but its consequences for health and socioeconomic outcomes remain uncertain. We examine these consequences in a fuzzy regression discontinuity design using electronic health records from the American Family Cohort (AFC) and longitudinal data from the Health and Retirement Survey (HRS). In AFC, we find that the diagnosis discontinuity peaks at 13 percentage points within six months and attenuates until it becomes insignificant at 42 months. We find a smaller and shorter-lived discontinuity for related prescriptions. These discontinuities remain across alternative specifications. Apart from specification and treatment window-dependent improvements in weight, BMI, and serum glucose, we do not observe a pattern of sustained improvement in cardiometabolic outcomes as a result of diagnosis and related care. In HRS, the threshold induces reductions of 22 percentage points in working for pay and 20.6 percentage points in labor-force participation at the two year follow-up. We do not observe a corresponding change in retirement, suggesting that diabetes diagnoses may lead to respondents reducing work without retirement. These reductions remain after several sensitivity checks but do not persist beyond two years. These findings suggest that health screening for chronic conditions may temporarily reduce labor supply, making employment responses relevant to the evaluation of broader costs and benefits of screening.

## 1. INTRODUCTION

Life-course transitions are increasingly recognized as critical determinants of socioeconomic well-being. Events such as getting married, losing a job, and graduating college can shape the social, economic, and psychological contours of people’s lives. Health can influence the timing and consequences of these transitions. A chronic condition may affect a person’s capacity for work, maintain relationships, and live independently. Receiving information about a chronic condition may influence the person’s behavior and the treatment they receive. Screening for health conditions may introduce both these changes: it can initiate clinical care and it can change the behavior of the bearers of the chronic condition. Understanding the consequences of screening requires examining how these changes interact to influence health and other life-course domains.

We examine these consequences using electronic health records from a large primary-care registry called the American Family Cohort (AFC) and the Health and Retirement Study (HRS) (1), a longitudinal survey of Americans aged fifty or older. In each dataset, we exploit a diabetes diagnostic threshold, using a fuzzy regression discontinuity design (RDD) to study the effects of treatment and outcomes. AFC allows us to characterize the clinical response at the treshold in terms of diagnosis and prescription and how this treatment affects cardiometabolic outcomes such as glucose homeostasis, lipids, body composition, care received, and others. Meanwhile, HRS allows us to study the effects on respondent health, their social lives, and how they engage with the labor market. Together, these analyses provide complementary evidence about the timing of care induced by the threshold and its consequences for the life course.

Due to its comorbidities, negative impact on energy metabolism, and prevalence in the population, type 2 diabetes (T2D) is an especially concerning disease. The prevalence of T2D is high and increasing for most age cohorts in the U.S. (2). The National Institute of Diabetes and Digestive and Kidney Diseases estimates that more than 40 million Americans have diabetes, of which approximately nine million are unaware of it. Its potential consequences extend across the life course and its symptoms and complications may alter the ability to participate in social and economic activities (3–5). Identifying previously undiagnosed diabetes creates an opportunity to address these risks, but the ultimate effects depend on the care and patient behavior that follows.

Treatment may affect lives through two broad biological pathways, as illustrated in Figure 1. The first is more short-term. Treating diabetes may improve insulin sensitivity and glycemic control, thereby improving focus, energy, metabolic health, and well being. This improvement in well being could have spillover effects onto other areas of one’s life: work, better relationships, and more ambition. The second area in which treating diabetes could affect the life course operates in the long term. Those with untreated diabetes have been shown to be at greater risk of developing serious complications, such as cardiovascular disease, vision loss, and severe neuropathy. These things could have a serious effect not only on one’s ability to work, but, in some cases, on one’s ability to live.

**Fig. 1.**
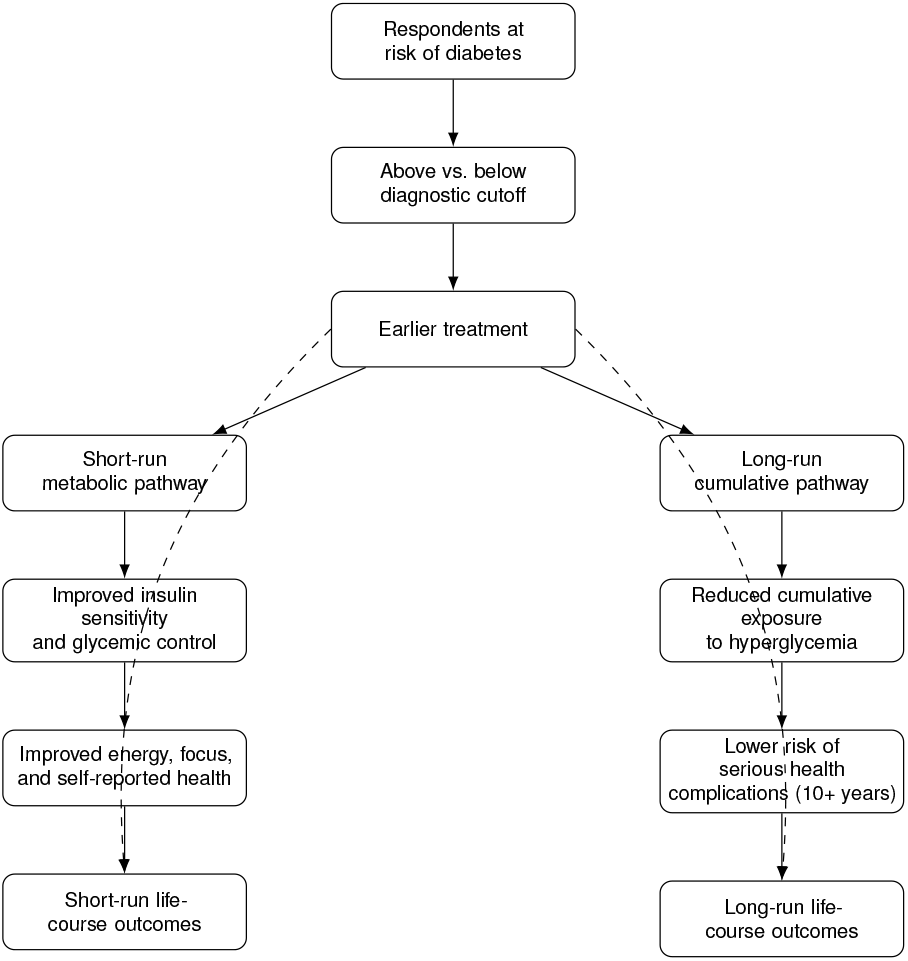
Conceptual pathways linking earlier diabetes treatment at the diagnostic cutoff to short-run and long-run life-course outcomes.

Much of the existing research on diabetes and socioeconomic outcomes compares people with a diabetes diagnosis with those without one, controlling for observed characteristics (6–10). Such comparisons provide helpful evidence but face two limitations. The first is that this research compares those with (likely treated) diabetes with a control group that likely does not have diabetes, whereas an equally interesting reference group may be those with biomarker-supported T2D that remains untreated. The second is that those diagnosed with diabetes likely have underlying risks and behaviors that observable characteristics simply cannot account for. Studies exploiting clinical thresholds offer the opportunity to examine the consequences of diagnosis and related care in patients with similar underlying risk (11, 12). Yet interpreting these estimates requires understanding what the clinical response tends to be and how long differences between the two sides of the threshold persists.

Glycated hemoglobin is a salient sign of the diabetes disease process (13). It is a continuous variable that correlates with glycemic exposure and clinical risk (14, 15). Clinical guidance from the American Diabetes Association (ADA) and the World Health Organization (WHO) uses an HbA1c level of 6.5 percent as a diagnostic threshold for diabetes. This guidance was established in 2010 (ADA) and 2011 (WHO). The ADA’s 2006 guidance recommended an HbA1c treatment goal below 7 percent for patients with diabetes (16). Because historical thresholds do not coincide with abrupt changes in underlying biological risk, the thresholds create a useful comparison for estimating the causal effects of diabetes diagnosis and treatment.

Using AFC data from 2012-2024, we follow clinical responses continuously after the initial HbA1c test. There is a range of actions at physicians’ disposal given an elevated HbA1c test. They may diagnose a patient with T2D, prescribe a medication, offer medical advice, or some combination of these. We estimate the discontinuities in diagnosis and prescriptions at the 6.5 percent threshold. We capitalize on the continuous clinical data and study how the responses vary according to the treatment window and according to treatment (i.e. diagnosis vs. prescription). We then analyze subsequent health outcomes using a fuzzy regression discontinuity design.

In HRS, we deploy a similar approach. The HRS began collecting dried blood spots (DBS) in 2006, prior to the passage of the ADA and WHO 6.5 percent guidance. At that point, the ADA’s guidance was to keep HbA1c below 7 percent. Consequently, HRS mailed respondents their blood sugar results with the 7 percent guidance stated next to the respondent’s results and encouraged the respondent to share the results with their physician. The letter also included a link to the ADA website. HRS used DBS every wave through 2016, so some of the follow-up waves occur after the change in ADA guidance. For this reason we use the 7 percent threshold as our primary focus, and report the results at the 6.5 percent threshold as secondary. The analysis in HRS complements what we study in AFC by exploring how the threshold induces change in health outcomes, social outcomes, and economic outcomes.

We find three main results. First, the AFC diagnosis discontinuity peaks at approximately 13 percentage points within six months of the initial test and attenuates thereafter, becoming statistically insignificant at 42 months. The diabetes medication discontinuity is smaller and shorter-lived. Second, although some specifications show early improvements in weight and BMI and lower glucose levels at later time periods, we do not observe a consistent pattern of improvement in measures of cardiometabolic health. Third, in HRS, crossing the 7 percent threshold is associated with reductions in employment. We find that the threshold induces a reduction in the likelihood of working by 22 percentage points and in labor force participation by 20.6 percentage points at the two-year follow-up. These results hold across several sensitivity checks, but do not persist in subsequent HRS waves. Retirement outcomes do not show a corresponding increase and imprecise first stage leaves the mechanism (i.e. diagnosis, prescription, or something else) unclear. Evaluating diabetes screening solely through health outcomes may overlook substantial economic effects, including adjustments to labor supply.

## 2. BACKGROUND

T2D typically develops when the body becomes insulin resistant, i.e., when cells stop responding normally to insulin to the point where the pancreas has to expand insulin production to maintain glucose homeostasis (17). These builders of insulin–beta cells– can compensate for a while by overproducing insulin, but lifestyle factors, environmental factors, genetics, and age can result in these cells ultimately being unable to keep up. T2D emerges when insulin demand exceeds beta-cell capacity to supply.

HbA1c has long been recognized as a valuable biomarker for tracking the development of and risks associated with T2D. Research found that complications of T2D, particularly the risk for diabetic retinopathy, started to become nonzero around 6.5 percent. Despite the recognition of its utility, however, HbA1c was not widely used for clinical diagnosis. The assays used to measure HbA1c showed substantial variation, to the point where its commercial values could not be relied on for diagnostics. But by the early 2000s, the variance across assays had fallen substantially, and in 2009 an international expert committee recommended that the 6.5 figure be used as a diagnostic threshold (18). The American Diabetes Association and the World Health Organization subsequently adopted it in their official guidance.

While HbA1c may be a diagnostic cut-off, it is not necessarily a biological cut-off. Some of the data shared by the international expert committee show a smooth curve around 6.5 percent (18). Subsequent research has shown that the value of HbA1c at which risk begins to increase varies by comorbidity, and that the risk for a given co-morbidity varies by race. Moreover, there is still some variance in HbA1c according to the assay (less than a few percentage points). These phenomena suggest that the care and attention received at an HbA1c in excess of 6.5 may not be connected with symptoms, and that symptoms may present at an HbA1c below 6.5. While the negative association between energy levels and HbA1c is strongest at levels of HbA1c much higher than 6.5, research shows that it is not uncommon to experience diabetes-related fatigue and other symptoms below 6.5 percent (19).

The literature has identified two broad pathways through which diabetes may impact the life course: 1) In the short-run, through diminished energy levels and related symptoms via impaired cellular glucose metabolism; and 2) A long-run pathway whereby more cumulative risk of more severe complications and comorbidities affect outcomes. These pathways identified in the research are illustrated in Figure 1.

## 3. DATA

Two primary types of data will be used in this paper: 1) electronic health records (EHR) and 2) panel dataset of households.

### Electronic Health Records

In this paper we use The American Family Cohort (AFC), a large, nation-wide dataset of primary care in the United States.^*^ AFC is populated from a registry of predominantly primary care clinics and covers interactions between more than 12,000 clinicians and approximately 8 million unique patients.

We restrict our sample to those patients who record a HbA1c test between 2012 and 2024 without a previous history of diabetes (type 1, type 2, or gestational diabetes). Patients with diagnosed prediabetes are not excluded. We select the first recorded HbA1c test, harmonizing across nine OMOP measurement concepts. We drop respondents whose first HbA1c test was recorded prior to 2012. We require that the observation period for the patients begin at least one year before the first test and continue for at least two years after. We include patients of all ages with the intention of later studying cohorts of different ages. A majority of patients AFC are age 50+, which means that whole-sample results in this paper will tend to reflect older Americans. These restrictions leave us with 662,663 patients in our eligible sample.

This sample has routine biometric and lab measurements done. Especially common measures include weight, height, blood pressure, glucose panels, and lipid panels. BMI is sometimes provided, but when absent we estimate it using the formula *BMI* = *weight/height*^2^.

AFC records information about the patient that we will use as control variables and heterogeneity analysis. These include sex, race, ethnicity, age, and U.S. state of care.

AFC has detailed data on prescriptions. In this study we focus on metformin, GLP-1 receptor agonists, SGLT2 inhibitors, sulfonylureas, DPP-4 inhibitors, thiazolidinediones, and insulin. In practice, however, when we are talking about a prescription we are referring to a metformin prescription. Metformin accounts for a vast majority of the prescriptions we see among the patients in our study. One reason is that we are focusing on patients without previous history of diabetes and those with HbA1c levels in the neighborhood of 6.5 percent. Insulin is rarely used for these cases. GLP-1 and SGLT2 drugs are newer, which may explain the relatively low incidence in our data.

### Panel Survey

For detailed coverage of the lives of older Americans, we use the Health and Retirement Survey (HRS). HRS is a longitudinal household panel of Americans over the age of 50. HRS welcomed its first cohort in 1992 and has followed that cohort until they leave the study or pass away. HRS adds new birth cohorts periodically, and interviews the spouses of the participants. In total, HRS interviews approximately 20,000 people each wave.

In 2006, HRS introduced dried blood spot (DBS) biomarkers–i.e. those taken from a few drops of blood from a finger or heel prick– that included HbA1c, but not other blood sugar measure. HRS randomly divided the sample into two groups that would alternate waves in which they would give DBS measurements. Consequently, respondents remaining in the sample have HbA1c measurements every four years through 2016.

HRS did not arrange consultations with physicians to discuss blood work results with the respondents, who instead received the result of their blood work within 4-6 weeks after the samples were drawn. Participants were encouraged to share the results with their doctor. Some results, such as blood pressure, were shared directly with the participant’s physician. HbA1c was not among the results shared directly with physicians. The letter^†^ contained the respondent’s HbA1c level along with guidelines from the American Diabetes Association regarding HbA1c levels that read (HRS included bolding), (20)

*‘****‘Hemoglobin A1c: X*.*X%***

*The hemoglobin A1c test shows the average amount of sugar in your blood over the last three months. The American Diabetes Association recommends that the goal of this result should be less than 7%. If your result is 7% or higher you should see your physician or other health professional to recheck this result and discuss with them*.

…

*American Diabetes Association http://www.diabetes.org”*

HRS used different labratories to handle the blood work over the years. These labs have assays that differ slightly from one another. HRS developed a way to map the values produced by the different laboratories to values that are compatible with those in the National Health and Nutrition Examination Survey (NHANES). Importantly, when HRS sent respondents the results of their blood work, they used the unadjusted laboratory readings, not the NHANES-equivalent values. They rounded the values to the nearest tenth decimal place. A detailed summary of the protocol HRS followed after the respondent blood draws can be found in Table A6.

In each survey wave 2006 to 2016, respondents were asked whether they had diabetes and whether they took diabetes medication orally or insulin.^‡^ These years also contain detailed information from different domains of the respondents lives, including anthropometrics, economic indicators, family structure, health care utilization, cognitive functioning, and measures of physical and mental health.

The availability of HbA1c measurements, diagnoses, prescription use, and detailed information about the respondents’ lives make the 2006 to 2016 a very useful starting point to build a cohort. In this study, we pool respondents that reported at least one HbA1c measurement. We take respondents first HbA1c value as “year 0”. We excluding respondents who reported having diabetes or taking diabetes medication by year 0. We then follow the respondents in the waves that follow from year 0. Pooling the respondents in this way increases our sample size significantly. Had HRS emailed resondents highlighting the 6.5 percent cut-off, using even a single year may have yielded enough observations for the present study. But the neighborhood of the 7 percent cut-off contains far fewer observation. With the pooled sample, we have approximately 1,500 respondents included in the selected bandwidth around 7 percent, including around 100 respondents to the right of the cut-off. We follow the respondents for four waves, or eight years, after the initial HbA1c test, at which point the number of observations begins to fall precipitously, mostly because we reached the most recent survey wave.

Together, AFC and HRS provide complementary evidence on diabetes treatment and the life course. AFC permits us to observe treatment continuously around the diagnostic threshold, while HRS provides repeated measures of health, social, economic, and financial outcomes for the same respondents as they move into retirement age.

## 4. EMPIRICAL STRATEGY

This paper capitalizes on the abrupt jump in the probability of treatment when HbA1c levels reach 6.5 to estimate the effects associated with common diabetes treatments. We utilize a Fuzzy Regression Discontinuity Design (RDD), which essentially uses the jump in probability at the cut-off as an instrumental variable for treatment:

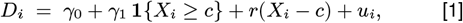

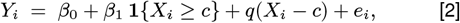

where *D*_*i*_ is the treatment status (in this setting, binary variables indicating diagnosis, prescription, either, or both) of individual *i*, **1** *{X*_*i*_ *≥c}* is an indicator function equal to one if the individual is at or above the cut-off *c*,^§^ *Y*_*i*_ is the outcome, *r*(*X*_*i*_ *−c*) and *q*(*X*_*i*_ *−c*) are nonparametric functions used to account for the nonlinear variation near the cut-off, and *u*_*i*_ and *e*_*i*_ are error terms. We allow *D*_*i*_ to reflect different types of treatment, such as having received a formal diagnosis of diabetes, having received prescription for diabetes, or both concurrently. The local average treatment effect for compliers at the cut-off is given by *τ* = *β*_1_*/γ*_1_ and is estimated via two-stage least squares.

A common analysis performed in RDD studies is to explore how *τ* changes over time. This analysis can be quite powerful, because the discontinuity identifies the causal effect and varying the time dimension gives insight into how long that effect persists. But the window of treatment matters as well. Mora and Rodríguez-Sánchez (2025) find that estimated reductions in direct healthcare costs become smaller as the delay between the laboratory test and diagnosis increases (21). Presumably, this attenuation is due to the fact that it decreases the time that observations are spent diagnosed.

We do not yet have a good understanding, however, of whether the discontinuity exists primarily for diagnosis or also for medication, and for which treatment windows the discontinuity exists. Many papers either use diagnosis or medication for the discontinuity, or create a variable equal to 1 if either is observed, which makes it difficult to attribute whether the ultimate effect on outcomes is due to diagnosis or prescription or a combination of the two.

This study attempts to clarify what exactly happens at the diagnostic threshold by comparing four different groups with the counterfactual of neither a diagnosis or a prescription: 1) Diagnosis only, 2) Medication only, 3) Either a diagnosis or a medication, and 4) Both a diagnosis and a medication. These treatments are distinct from one another, such that those in group 1 are not in group 2 or group 4. We vary the treatment windows to see how long the discontinuities in treatment remain. This exercise will yield important evidence for future diabetes RDD studies in the United States of what treatments are becoming more likely at the cut-off and the timing of these treatments. It builds on the work of Alalouf, Miller, and Wherry (2024), who (11)

The empirical strategy shown in Equation 1 and Equation 2 is made possible by the AFC data because the treatment is continuously observed, whether it occurs the same day as the test or one year later. Despite all of the advantages of HRS—such as the breadth of information collected about the respondents—it has the disadvantage that the treatment is not continuously observed.

In HRS, respondents are interviewed every two years (or around 100 weeks after receiving the letter with their HbA1c results). In the results section, we will present the first stage results for HRS data using Equation 1, but because the treatment window is wide, we anticipate that the first stage instruments will be weak. As such, we report the results for Equation 2 as our primary specification. The reduced-form jump in the outcome identifies the local intent-to-treat effect of crossing the cut-off when the outcome would otherwise be continuous at the cut-off (22). This differs from the effect of diagnosis among patients at the cut-off whose diagnosis is induced by crossing it, an effect commonly referred to as the local average treatment effect or LATE (23). Under the usual fuzzy-RDD assumptions, the reduced-form effect equals the LATE multiplied by the first-stage jump in the probability of diagnosis and is therefore smaller in magnitude when that jump is between zero and one.

Because the HbA1c measurements sent to the respondents were rounded to the nearest tenth decimal place, we round to the same place. This creates a continuous variable that has mass points at integers. We therefore adjust for mass points in all regressions, adjusting the optimal bandwidth selection to ensure sufficient support on either side of the cut-off. We use triangular kernel weights to prioritize observations closer to the cut-off. Standard local-polynomial RDD uses these kernel weights; unit-specific survey weights are an optional additional multiplier (24). In all RDD models we report bias-corrected point estimates and standard errors, and correct for heteroskedasticity in the standard errors.

### Ensuring sample attenuation and bandwidth selection do not drive results

Because we use an optimal bandwidth procedure, the bandwidth may change depending on what data are available. Sample attenuation–i.e. patients no longer receiving care or being observed in the data–could make results even more susceptible to bandwidth selection and are a concern themselves. To ensure that our first stage results are not being driven by merely by these factors, we run a robustness check by setting the bandwidth at the 6-month treatment window and use that same bandwidth for every other treatment window. In so doing, we only use patients that are observed during the full treatment window (i.e. 96 months). We report these figures in the annex.

## 5. RESULTS

### Electronic Health Records

We begin by examining the the discontinuities in treatment. In Figure A2 through Figure A9, we plot the results for Equation 1, the first-stage, while varying the treatment window. Each figure represents a different subsample, and each quadrant within each figure shows the first stage effect for the four potential treatments: a) diabetes diagnosis, b) diabetes prescription, c) either a diagnosis or a prescription, or d) both a diagnosis and a prescription. The plots generally show an attenuation over time, reflecting patients just under the threshold eventually receiving a diagnosis or a prescription.Figure A2 shows estimates for the entire sample. The strength of the first stage for diagnosis peaks at around 13 percentage points at a treatment window of 6 months. A jump of 13 percentage points is a massive increase. The probability of diagnosis below the cut-off is around 10 percent, so this represents a more than doubling of the diagnosis rate (Figure 2 illustrates this well). The jump in the probability of a diabetes prescription is positive and statistically significant for whole sample within the first six months, but this jump is modest its statistical significance is notably weaker than with the discontinuity in diagnosis. Figure A3 through Figure A7 demonstrate that the cut-off is robust across genders and race. Figure A8 does not show a statistically significant discontinuity in either treatment for young adults. The confidence intervals for young adults are especially wide. For these first stage results, we use the optimal bandwidth from the model with threatment window of 6 months for each of the other months. This will complement our first-stage reporting of the fuzzy IV results, whose bandwidth is dynamically determined at each step. These figures here show that the results observed in the full model are not derived from the bandwidth changing, introducing a different population that potentially correlates with the outcomes. The selected bandwidths for Figure A2 through Figure A9 are displayed in Table A3

Figure A9 shows that the discontinuity is strong, especially for diagnosis, in adults age 50+, the same demographic as our HRS cohort. That the results for age 50+ in AFC mirror those of the whole sample is unsurprising, since a majority of patients in AFC are age 50+. The key here is that the same demographic of which HRS is nationally representative shows a strong discontinuity in diabetes diagnosis in the AFC data. The window appears statistically significant through 36 months after the initial HbA1c test (Panel A). Diabetes-related prescriptions show a smaller discontinuity: The increase in the likelihood of prescription is only half as high and the treatment window is only about 6 months. This result should color the interpretation of the second stage results. Any results we see there, then, are predominantly attributable to the diabetes diagnosis–unless, of course, we think that a few months head start on a metformin prescription is what is driving the results.

**Fig. 2.**
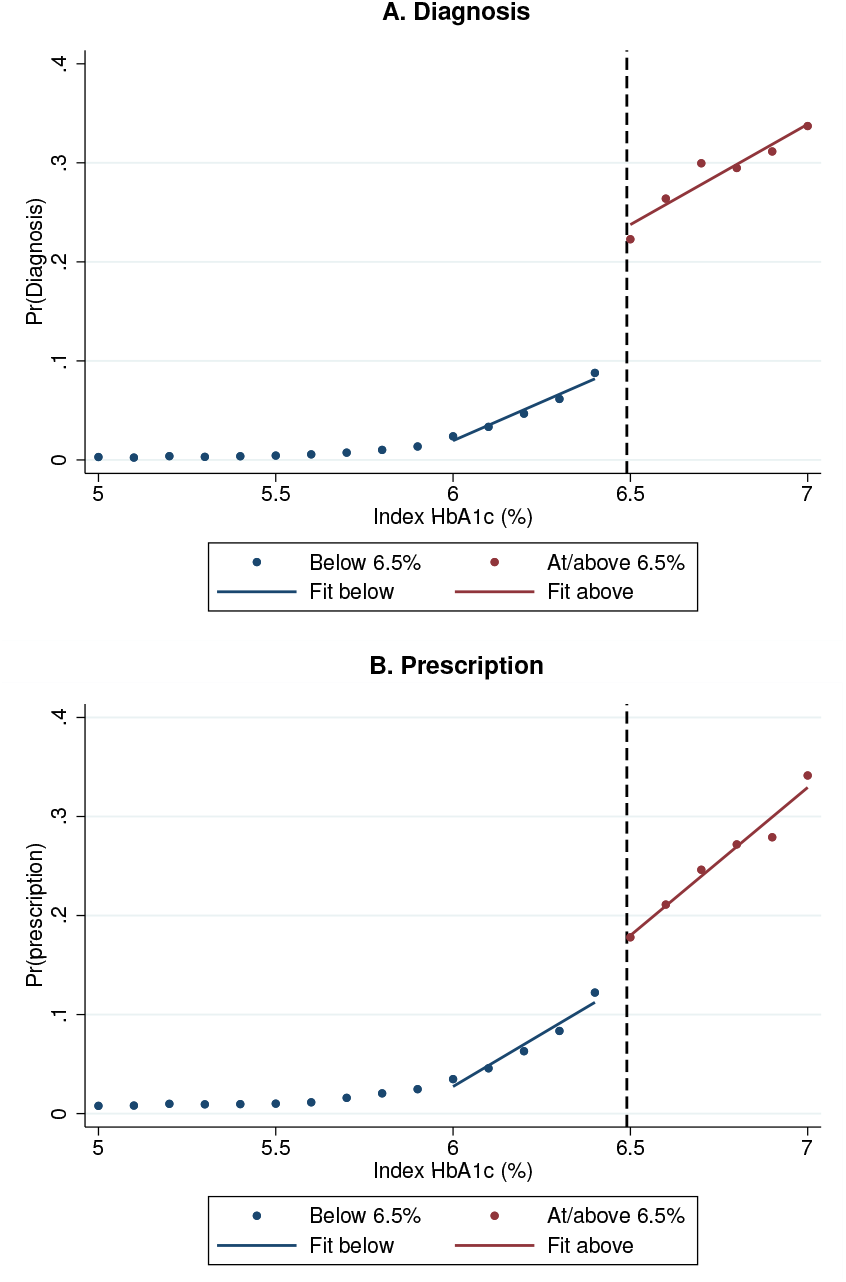
American Family Cohort: Discontinuities in the probabilities of diagnosis and prescription at HbA1c = 6.5. *Note*: Points show binned means of the relevant treatment indicator around the HbA1c diagnostic cut-off of 6.5%. Solid lines show fitted values estimated separately on either side of the cut-off using the first-stage specification in Equation 1, where *D*_*i*_ is defined as diagnosis in Panel A and prescription in Panel B. The sample includes AFC patients with an HbA1c test between 2012 and 2024 and no previous diabetes history.

Next we report the fuzzy RDD results on cardiometabolic outcomes and care. Figure 3, Figure 4, and Figure 5 depict the second-stage estimates for diagnosis, either, and both respectively. These figures shows the effect on the outcome varies across time (“outcome window” in the x-axis) and according to treatment window (color coded and in the legend). Each figure shows this effect for eight outcomes: 1) HbA1c level, 2) serum glucose, 3) BMI, 4) weight, 5) systolic blood pressure, 6) diastolic blood pressure, 7) LDL cholesterol, and 8) Number of HbA1c tests. Specifications with a first stage wald statistic^¶^ of less than 10 are not displayed. The corresponding figure containing the prescription results is not displayed because ony four of the 144 tests have a strong enough first stage. A full table of first stage Wald statistics can be found in Table A4.

**Fig. 3.**
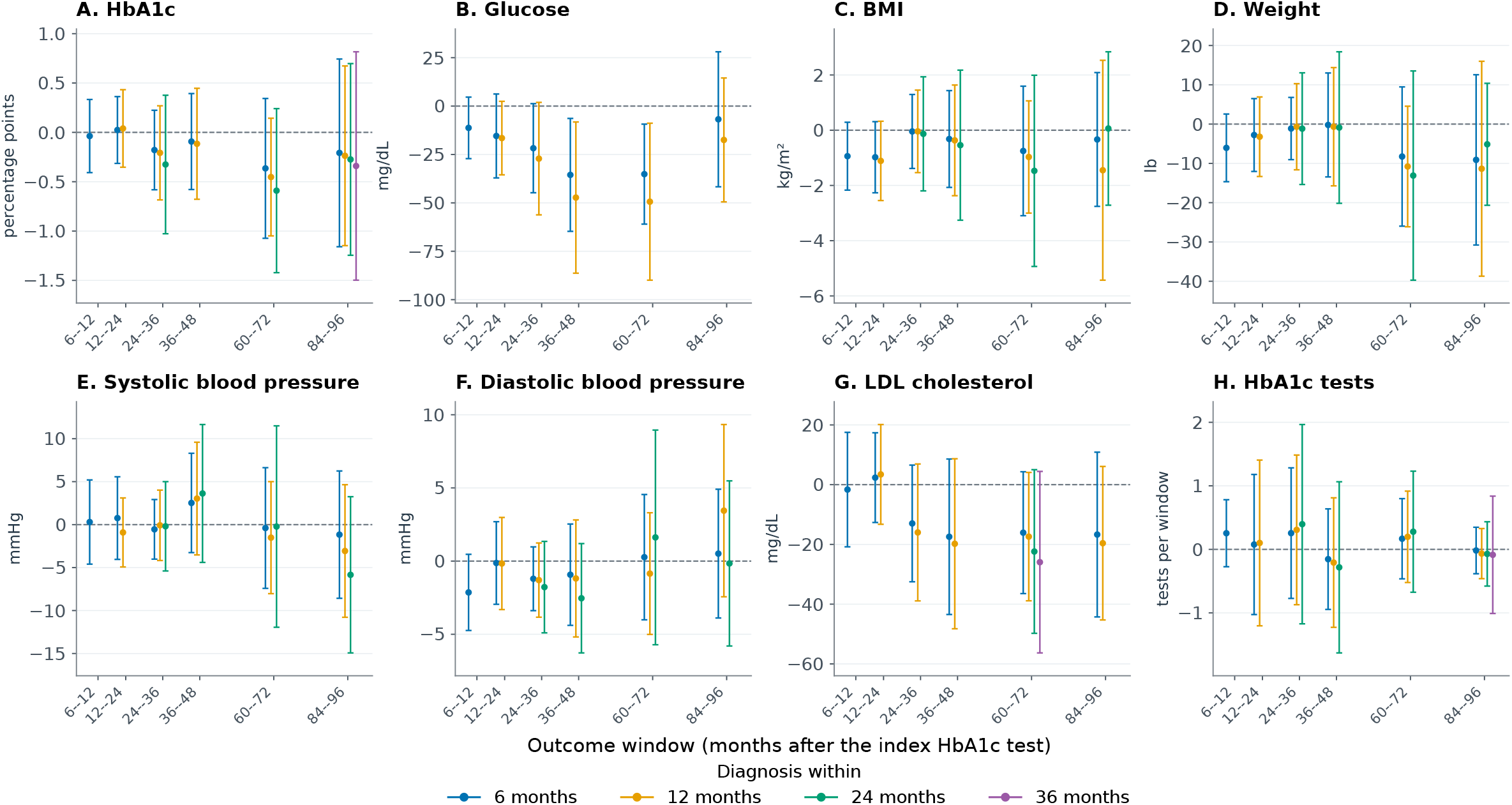
Subsequent health outcomes and monitoring by timing of diabetes diagnosis. *Note*: Points show bias-corrected fuzzy regression-discontinuity estimates at the HbA1c threshold of 6.5%; capped vertical lines show robust 95% confidence intervals. Treatment is defined as a recorded diabetes diagnosis within 6, 12, 24, or 36 months after the index HbA1c test, shown from left to right within each outcome window in blue, gold, green, and purple. The horizontal axis measures time from the index test, not from treatment; small horizontal offsets distinguish treatment horizons. Combinations in which the treatment window overlaps the outcome window are omitted. Each cell uses a local-linear specification with local-quadratic bias correction, triangular kernel weights, MSE-selected bandwidths, mass-point adjustment, controls for sex, race, ethnicity, state, index year, age, and age squared, plus the outcome’s baseline value and a baseline-missing indicator when available; standard errors are clustered by care site. The available-follow-up sample and estimation bandwidths vary by outcome and timing combination. Estimates are in the units shown on each panel. HbA1c test counts refer to the full outcome window: the first window spans six months and the remaining windows span twelve months. Estimates and their intervals are omitted when the squared robust first-stage *z* statistic is below 10; this screen matches the weak-first-stage flags for these outcomes. This statistic is not a Kleibergen–Paap F statistic, and the screen does not guarantee strong identification. Of 144 temporally valid estimates, 115 are displayed and 29 are omitted because of weak first stages. Diagnosis estimates may include changes in medication and other care following diagnosis.

**Fig. 4.**
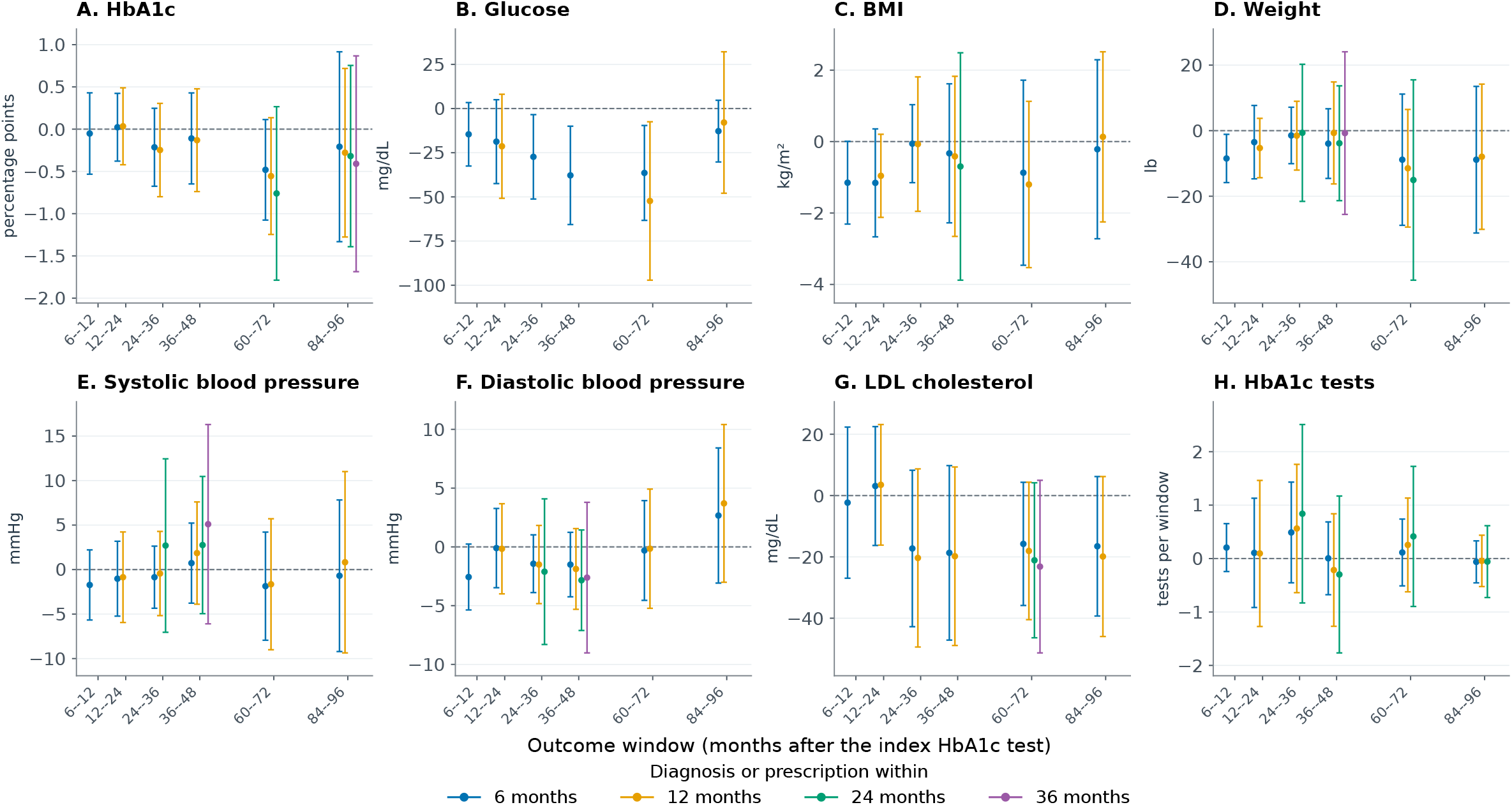
Subsequent health outcomes and monitoring by timing of either diabetes diagnosis or medication prescription. *Note*: Points show bias-corrected fuzzy regression-discontinuity estimates at the HbA1c threshold of 6.5%; capped vertical lines show robust 95% confidence intervals. Treatment is defined as either a recorded diabetes diagnosis or a diabetes-medication prescription within 6, 12, 24, or 36 months after the index HbA1c test, shown from left to right within each outcome window in blue, gold, green, and purple. The horizontal axis measures time from the index test, not from treatment; small horizontal offsets distinguish treatment horizons. Combinations in which the treatment window overlaps the outcome window are omitted. Each cell uses a local-linear specification with local-quadratic bias correction, triangular kernel weights, MSE-selected bandwidths, mass-point adjustment, controls for sex, race, ethnicity, state, index year, age, and age squared, plus the outcome’s baseline value and a baseline-missing indicator when available; standard errors are clustered by care site. The available-follow-up sample and estimation bandwidths vary by outcome and timing combination. Estimates are in the units shown on each panel. HbA1c test counts refer to the full outcome window: the first window spans six months and the remaining windows span twelve months. Estimates and their intervals are omitted when the squared robust first-stage *z* statistic is below 10; this screen matches the weak-first-stage flags for these outcomes. This statistic is not a Kleibergen–Paap F statistic, and the screen does not guarantee strong identification. Of 144 temporally valid estimates, 106 are displayed and 38 are omitted because of weak first stages. The treatment indicator turns on when either event is recorded, so estimates combine patients induced across the threshold into diagnosis, prescription, or both.

**Fig. 5.**
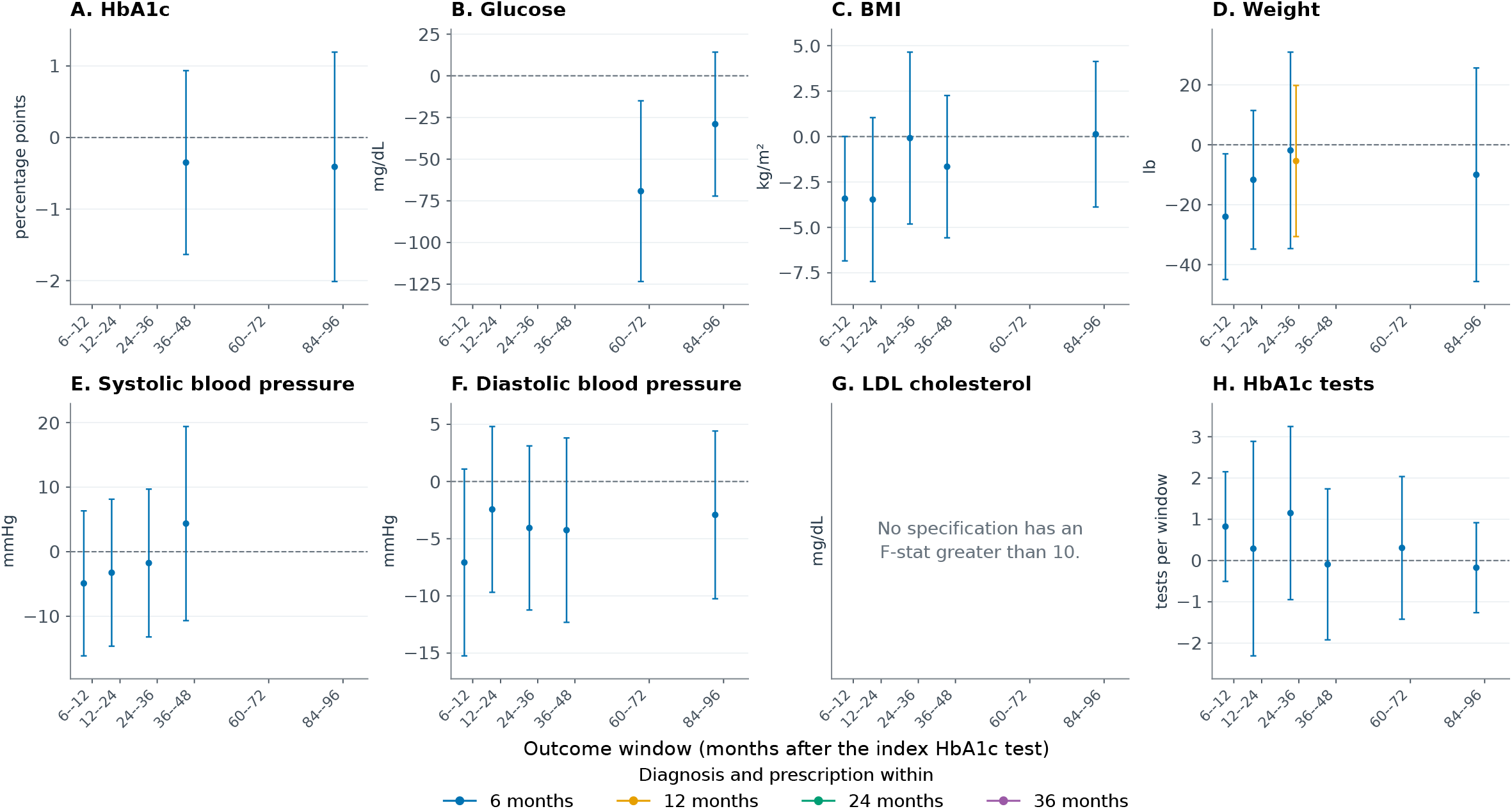
Subsequent health outcomes and monitoring by timing of both diabetes diagnosis and medication prescription. *Note*: Points show bias-corrected fuzzy regression-discontinuity estimates at the HbA1c threshold of 6.5%; capped vertical lines show robust 95% confidence intervals. Treatment is defined as both a recorded diabetes diagnosis and a diabetes-medication prescription within 6, 12, 24, or 36 months after the index HbA1c test, shown from left to right within each outcome window in blue, gold, green, and purple. The horizontal axis measures time from the index test, not from treatment; small horizontal offsets distinguish treatment horizons. Combinations in which the treatment window overlaps the outcome window are omitted. Each cell uses a local-linear specification with local-quadratic bias correction, triangular kernel weights, MSE-selected bandwidths, mass-point adjustment, controls for sex, race, ethnicity, state, index year, age, and age squared, plus the outcome’s baseline value and a baseline-missing indicator when available; standard errors are clustered by care site. The available-follow-up sample and estimation bandwidths vary by outcome and timing combination. Estimates are in the units shown on each panel. HbA1c test counts refer to the full outcome window: the first window spans six months and the remaining windows span twelve months. Estimates and their intervals are omitted when the squared robust first-stage *z* statistic is below 10; this screen matches the weak-first-stage flags for these outcomes. This statistic is not a Kleibergen–Paap F statistic, and the screen does not guarantee strong identification. Of 144 temporally valid estimates, 29 are displayed and 115 are omitted because of weak first stages. No LDL cholesterol estimates pass the first-stage screen. The treatment indicator turns on only after both events are recorded.

A key outcome is that diagnosis does not have a statistically significant effect on the primary marker of disease pregression (Panel A of Figure 3). It also does not seem to have an effect on the number of HbA1c tests undertaken (Panel H). While these outcomes are measured with less precision without baseline outcomes as controls, the estimated coefficients are quite close to 0, suggesting a lack of precision is not behind these null results.

Panel B of Figure 3 suggests that there might be a longer-term effect of diagnosis on glucose approximately 3 to 6 years after the initial test. This effect is meaningful at around 30 mg/dL. This effect is seen in the “either” treatment Figure 4) and in one test in the “both” treatment Figure 5. The effect tied to diagnosis is seen in both the 6-month and 12-month treatment window. Both the either treatment and the both treatment show a statistically significant decrease in BMI and weight 6-12 months after the initial tests, but this effect does not persist after 12 months and is only present for the 6 months treatment window. Across all three figures, the treaments generally seem to have effects that are directionally consistent with improvements cardiometabolic health (e.g. HbA1c, glucose, BMI, weight, blood pressure, cholesterol) and an increase in care received (i.e. number of HbA1c tests)—but are generally not statistically significant. One possible interpretation of these results is that in the real world a diabetes diagnosis may have a small influence on biological measures of health but these effects are lost in the noise created by significantly more influential forces affecting these measures.

### Health and Retirement Study Cohort

As HRS respondents are sent a letter highlighting an HbA1c level of 7 percent as the clinical threshold, our main results here focus on this threshold. We generally do not see very precise results at the first stage. Receiving a diagnosis has a standard error of 0.096, and receiving a prescription has a standard error of 0.082 (column 2 of Table A11). These values are observed two years after the respondents’ first HbA1c test in HRS. This is plausibly consistent with what we see in respondents age 50+ (Figure A9), where the effect has already attenuated by 24 months and is statistically insignificant by 3.5 years. We therefore present here reduced form results that capture the effect induced at the threshold. The effective changes that likely to occur at the threshold are an early diagnosis and perhaps a slight head start on metformin, though by presenting the reduced form results it is impossible to attribute any effects to one or the other.

We find that crossing the threshold leads to disruptions in the labor market two years after the initial HbA1c test (column 1 of Table 1). Those just above the cut-off saw a 22 percentage point reduction in the likelihood of reporting any work and a 20.6 percentage point reduction in the probability in being in the labor force. There is some evidence that respondents reduced hours worked by about 5.7 hours per week, but this is marginally significant. The point estimate on log of earnings is negative but statistical insignificant. The negative effects on employment and retirement observed at the two-year follow-up are not observed in subsequent follow-ups. Indeed, the point estimates are close to zero. This suggests that the negative effect on employment is transient.

**Table 1.** Health and Retirement Study reduced-form results at the 7.0% cutoff.

| Outcome | 2 years |  | 4 years |  | 6 years |  | 8 years |  |
| --- | --- | --- | --- | --- | --- | --- | --- | --- |
|  | (1) |  | (2) |  | (3) |  | (4) |  |
| | $\hat{\beta}$<br>(SE) | $h$<br>$N_{\text{eff}}$ | $\hat{\beta}$<br>(SE) | $h$<br>$N_{\text{eff}}$ | $\hat{\beta}$<br>(SE) | $h$<br>$N_{\text{eff}}$ | $\hat{\beta}$<br>(SE) | $h$<br>$N_{\text{eff}}$ |
| Completely retired | **-0.094\***<br>(0.054) | 1.061<br>1,680/97 | 0.022<br>(0.071) | 0.946<br>1,093/77 | 0.061<br>(0.094) | 1.338<br>2,950/79 | 0.005<br>(0.063) | 0.995<br>816/61 |
| Retired in some capacity | 0.086<br>(0.069) | 1.559<br>6,025/116 | 0.057<br>(0.077) | 1.108<br>1,991/85 | 0.100<br>(0.121) | 1.067<br>1,307/69 | 0.030<br>(0.088) | 1.004<br>1,138/63 |
| Working | **-0.220\*\*\***<br>(0.074) | 0.996<br>1,209/93 | 0.079<br>(0.073) | 0.734<br>533/70 | -0.024<br>(0.095) | 1.306<br>2,943/79 | -0.029<br>(0.050) | 0.713<br>428/56 |
| Hours worked | **-5.663\***<br>(3.424) | 1.197<br>2,193/103 | 5.131<br>(3.707) | 0.652<br>361/67 | -4.642<br>(3.966) | 1.267<br>2,215/74 | -1.734<br>(2.359) | 0.622<br>285/52 |
| In the labor force | **-0.206\*\*\***<br>(0.070) | 1.127<br>2,206/103 | 0.043<br>(0.062) | 0.768<br>520/70 | -0.107<br>(0.104) | 1.091<br>1,260/67 | **0.099\*\***<br>(0.050) | 1.305<br>2,419/68 |
| Not working and not retired | 0.001<br>(0.066) | 1.153<br>2,234/105 | -0.084<br>(0.073) | 0.875<br>754/71 | 0.044<br>(0.064) | 1.266<br>2,265/76 | -0.021<br>(0.068) | 1.224<br>1,950/67 |
| Not in the labor force & not retired | 0.019<br>(0.062) | 1.229<br>2,942/108 | -0.011<br>(0.058) | 1.056<br>1,512/79 | 0.053<br>(0.062) | 1.647<br>5,315/85 | -0.009<br>(0.066) | 1.276<br>1,950/67 |
| Limited by health | 0.052<br>(0.081) | 1.018<br>1,494/87 | 0.068<br>(0.083) | 1.344<br>3,111/80 | -0.103<br>(0.083) | 1.212<br>2,039/71 | 0.104<br>(0.119) | 0.915<br>740/55 |
| Log earnings | -0.401<br>(0.866) | 1.032<br>1,680/97 | **-1.512\***<br>(0.859) | 1.286<br>2,624/88 | -1.269<br>(0.887) | 1.407<br>3,711/81 | -0.223<br>(0.761) | 1.067<br>1,138/63 |
| Receives SSDI benefits | 0.015<br>(0.049) | 1.061<br>1,680/97 | -0.015<br>(0.046) | 0.708<br>535/70 | 0.009<br>(0.016) | 0.701<br>478/62 | 0.024<br>(0.045) | 0.963<br>816/61 |
| Log SSDI benefits | 0.183<br>(0.650) | 1.069<br>1,680/97 | -0.125<br>(0.540) | 1.275<br>2,624/88 | -0.314<br>(0.489) | 1.131<br>1,734/74 | -0.189<br>(0.414) | 1.151<br>1,496/66 |
| Basic physical self-care/functioning | -0.067<br>(0.225) | 1.181<br>2,234/105 | **-0.335\*\***<br>(0.158) | 0.931<br>1,093/77 | 0.010<br>(0.228) | 1.185<br>1,733/74 | -0.251<br>(0.286) | 0.966<br>813/61 |
| Independent everyday functioning | -0.176<br>(0.164) | 0.953<br>1,209/93 | -0.236<br>(0.157) | 0.993<br>1,093/77 | 0.093<br>(0.169) | 0.980<br>942/67 | -0.129<br>(0.240) | 0.973<br>813/61 |
| Self-reported health | 0.165<br>(0.169) | 1.025<br>1,679/97 | -0.101<br>(0.215) | 1.232<br>2,622/88 | 0.088<br>(0.201) | 1.205<br>2,260/76 | -0.253<br>(0.160) | 1.204<br>1,947/67 |
| Cognitive score | 0.335<br>(0.682) | 1.164<br>2,167/101 | -0.115<br>(0.650) | 1.437<br>4,214/89 | -0.362<br>(0.869) | 1.217<br>2,159/71 | 0.169<br>(0.740) | 1.111<br>1,419/62 |
| Days hospitalized the past 2 years | 0.067<br>(0.203) | 1.184<br>2,221/103 | -0.427<br>(0.305) | 0.729<br>527/69 | **0.906\*\***<br>(0.457) | 1.198<br>1,713/74 | -0.503<br>(0.369) | 0.920<br>803/61 |
| Married | 0.061<br>(0.056) | 1.070<br>1,680/97 | 0.034<br>(0.030) | 1.079<br>1,512/79 | -0.045<br>(0.055) | 1.609<br>5,315/85 | 0.095<br>(0.066) | 1.151<br>1,496/66 |

*Note:* This table reports bias-corrected reduced-form local-linear regression discontinuity estimates for Health and Retirement Study respondents aged 50 and older with no prior diabetes diagnosis or diabetes medication use. Robust standard errors appear in parentheses. $h$ is the estimation bandwidth, the same on both sides of the 7.0% HbA1c cutoff; $N_{\text{eff}}$ lists effective observations below/above the cutoff. Regressions use triangular kernel weights and adjust for mass points in the running variable. Covariates include the polygenic score for HbA1c, an indicator for missing genetic data, age at first test and its square, indicators for whether the respondent's father and mother are alive, and the natural logarithm of baseline earnings plus one, which includes respondents with zero earnings. \* $p < 0.10$ , \*\* $p < 0.05$ , and \*\*\* $p < 0.01$ , based on bias-corrected $p$ -values. The displayed basic physical self-care/functioning outcome comes from the ADL5A variable and measures activities such as bathing, dressing, eating, walking across a room, and getting in or out of bed. The independent everyday functioning outcome comes from the IADL5A variable and measures activities such as managing medications and finances and shopping.

The first two rows in Table 1 show the effects on whether the respondent is completely retired or retired in some capacity (either partially or completely). There is a slight reduction in the likelihood of being completely retired at year two, but it is marginally significant. The point estimate for any retirement is slightly positive, suggesting that there may be a small increase in the likelihood of being partially retired. In any case, this effect is equally transient and does not last beyond year two. There are some isolated effects effects with a moderate amount of statistical significance at different years (e.g. reduced difficulties reported basic physical functioning), but it does not fit neatly with other results, so we hesitate to read too much into them.

Some of the observed effects at year two are quite large. Around 48 percent of respondents were working for pay at baseline (column 1 of Table A1), so a 22 percent change represents a 45 percent change. We discuss this more in section 6.

## 6. ROBUSTNESS

We stress test the fuzzy RDD for AFC cardiometabolic outcomes in a number of ways. First, we jitter the bandwidth, decreasing it by 25 percent, increasing it by 25 percent, and increasing it by 50 percent. Second, we run two additional models with different sets of controls. One has only the baseline value of the outcome (except for HbA1c and HbA1c tests, for which we take respondents first test), and the other has no control variables at all. Third, we allow for the data on either side of the cut-off to be fit with a second-order polynomial. The RDD literature normally fits the data with a first-order or linear model. And finally, we drop observations that are exactly equal to the cut-off, in case some randomness at the cut-off is driving the results. We run these robustness checks for each of our four treatments: diagnosis, medication, either, or both. We run the robustness checks for outcomes 12-24 months after the initial HbA1c test. We do not display the estimates for models that do not pass the first stage (i.e., their first-stage robust Wald statistic is less than ten). The full results of these robustness checks can be viewed in Figure A10 through Figure A13 with the original point estimate and confidence interval included for comparison.

The robustness checks consistently agree with the main specification 12-24 months after the initial test. In fact, we do not observe a single robustness check result that is discordant with the main estimate. These robustness checks reinforce the idea that primary care diagnosis of diabetes in the real world does not result in significant changes to a wide range of cardio-metabolic outcomes.

There are several important sensitivity checks that we do with our panel data RDD model. First, we run the model with baseline values of the outcomes instead of the outcomes themselves. Second, we run a model that only uses baseline values of the outcomes as control variables. Third, we run a model that includes no control variables, including baseline outcome values. Fourth, we exclude observations that are exactly equal to the cut-off. Fifth, we jitter the bandwidth, reporting the results at 25 percent smaller bandwidth and 25 percent larger bandwidth. And sixth, we report discontinuities of control variables at the cut-off.

We also report results at the 6.5 percent cut-off. Because the respondents were sent a letter with the explicit 7 percent threshold right next to their HbA1c test result, we believe that the 7 percent is a cleaner cut-off than 6.5 percent. That said, it is possible that engaged respondents could have searched the internet to assist in interpreting their results or followed the letter’s link to the American Diabetes Association (ADA) website, encountered the 6.5 percent cut-off, and interpreted their results in light of it. Moreover, it is also possible that they heeded HRS’s encouragement to share the test results with their doctor. And it is possible that the engaged respondent or the respondent’s doctor may have anchored the result with the prevailing 6.5 percent ADA threshold. We would anticipate the jump at this threshold, should it exist, to be significantly weaker than the 7 percent threshold. We nonetheless run all of the same tests for the 6.5 percent cut-off that we do for 7 percent cut-off, including all robustness checks, to give a complete picture. We also run an additional robustness check where we ensure that the bandwidth does not include the other cut-off.

The robustness checks at the 7 percent cut-off generally support the results of the main specification that cut-off induces a greater likelihood to leave the labor force, some of whom decide to retire (Table A13. Some precision is lost when excluding control variables, especially the outcome at baseline (column 3 of Table A13). The effects attenuate slightly when respondents at the seven percent cut-off are excluded (column 4 of Table A13), but this could also be that we are removing the largest, most heavily-weighted bucket on the right side of the cut-off when there are only around 100 observations there to begin with.

The 6.5 percent cut-off says tells a different, albeit less consistent, story than is seen with the seven percent cut-off. The cut-off induces an increase in the likelihood of being in the labor force and working for pay, which are both significant at the 5 percent level (column 1 of Table A7). There is no effect on the measures of retirement. The effect on wages is positive, mirroring the employment effects, but not statistically significant. We had concerns with the 6.5 threshold *ex ante*, and there are also concerns with interpreting results at this threshold *ex post*. The conclusions are not stable across the robustness checks shown in Table A12. Excluding all covariates cuts the coefficient on working in half and makes both the working and labor force results insignificant. Excluding the cut-off causes both effects to be insignificant. The results have limited statistical significance and variable point estimates when changing the bandwidths. And perhaps most importantly, both of these effects disappear when we used a balance panel that reports in all four follow-up waves (shown in Table A8). For the aforementioned ex ante and ex post reasons, we therefore report the results at the 7 percent threshold as our primary results, results which demonstrate greater stability across specifications.

## 7. CONCLUSION

Clinical thresholds can shape the patients receive even while underlying biology changes smoothly around the threshold. In this paper, we examine the consequences of these thresholds using a large primary care electronic health record and a longitudinal panel. The American Family Cohort (AFC) allows us to examine the discontinuities that exist at the hemoglobin A1c (HbA1c) threshold of 6.5 percent for diabetes diagnosis and diabetes-related prescriptions. We use the Health and Retirement Study to examine how socioeconomic outcomes among older adults in the U.S. changes around the 7 percent cut-off recorded in letters sent to the patients and very well their physicians. These analyses together reveal the clinical responses generated by the threshold and how they in turn may affect socioeconomic outcomes.

The AFC results show that the threshold generates a substantial discontinuity in the form of diabetes diagnosis, but a smaller and less persistent discontinuity for prescriptions. We study how this effect varies with the treatment window. The diagnosis effect peaks at around 13 percentage points when diagnosis is measured within six months following the first HbA1c test and attenuates as the treatment window widens, becoming indistinguishable from 0 at around 42 months (or 3.5 years). This finding holds even when we freeze the follow-up sample and bandwidths. Meanwhile the effect on prescription peaks at around 7 percentage points, but becomes insignificant after just one year. The timing and duration of the discontinuity therefore matter when interpreting subsequent outcomes.

While the discontinuities suggest earlier diagnosis, the diagnosis and any related prescriptions do not produce patterns of sustained improvements across a range of common cardiometabolic outcomes. While we find some evidence of early reductions in BMI and weight and improvements in later measures of serum glucose, these outcomes vary across outcomes, treatment windows, and specifications that do not give us durable confidence in these results. A common pattern is to see point estimates in the direction of improvement but with wide confidence intervals, a finding that is potentially consistent with the story of modest improvements in outcomes that are ultimately affected by more powerful forces. Importantly, we do not see a diabetes diagnosis leading to improvements in the primary marker of diabetes, HbA1c, or related care, proxied by the number of HbA1c tests. We are not able to identify the effect of prescriptions on cardiometabolic outcomes, as the specifications often have a weak first stage. From the AFC analysis we conclude that changes in perceive health do not necessarily lead to broad changes in health.

In HRS, we find significant effects on employment at the year two follow-up. Crossing the 7 percent threshold corresponds to a 22 percentage point reduction in the probability of working for pay and a 20.6 percentage point reduction in labor-force participation. These results hold when examining a balanced panel with data for all five waves and across several alternative specifications. These effects do not show up in later HRS waves, suggesting the employment effect lasts less than four years. While we find a negative impact on employment, we do not find a positive effect on retirement, suggesting that the respondents may be taking a break from work with the intention of working at some point in the future. We know from the AFC data that the biggest change to people’s lives at the threshold is increased likelihood of diagnosis. We do not see evidence that diabetes diagnosis or a related prescription leads to significant changes in underlying biology, a finding which agrees with previous literature ((11)). In the HRS data, there may be some evidence of increased retirement, but the results are inconsistent and even if significant would not be large enough to explain the negative effects on employment. In the face of this evidence, it seems that a diabetes diagnosis itself, rather than the underlying disease, leads to respondents to take a break from the labor force. That treating chronic disease might influence labor supply has important implications for health care policy, especially in a system where health insurance, and thereby continuity of care, is often tied to employment.

Our findings are qualified by several limitations. The HRS estimates rely on relatively few observations above the threshold. Some baseline continuity and placebo checks also show discontinuities, leaving concerns that our sensitivity checks do not fully resolve. While we do our best to connect the relevance of AFC to HRS, differences in populations, thresholds, and clinical settings limit how relevant the connection can be. Ultimately, the estimates therein apply locally to respondents near the thresholds.

The conclusions of this study have implications for both the evaluation of diabetes screening and the use of clinical thresholds in causal research. Threshold studies should establish which responses are induced and the treatment windows for which the threshold exists. Future work, wherever possible, should examine socioeconomic outcomes in addition to biological outcomes to understand the comprehensive effect of medical information. Datasets with more frequent observation of health events and work events could shed light on the mechanisms, timing, and heterogeneous effects of the findings in this study.

## Data Availability

The data that support the findings of this study are derived from the American Family Cohort (AFC), a proprietary electronic health record database containing patient-level health information. The underlying data are not publicly available because of privacy, ethical, and licensing restrictions. Researchers may obtain access to AFC data through the appropriate application and approval processes established by the data custodians. Researchers may obtain access to restricted and senstivite HRS data through appropriate application and approval processes established by the data custodians.

https://med.stanford.edu/phs/data/american-family-cohort--afc-.html

https://hrs.isr.umich.edu/data-products/restricted-data

## Acknowledgments

The Health and Retirement Study (HRS) is sponsored by the National Institute on Aging (grant number U01AG009740) and is conducted by the University of Michigan. This study uses restricted-access HRS data, including biomarkers derived from dried blood spot (DBS) samples and polygenic scores (PGS), obtained under an HRS Restricted Data Access Agreement. We acknowledge the University of Michigan and HRS for providing access to these data. The statistical outputs derived from restricted HRS data and presented in this paper underwent the HRS disclosure review process and were approved for public release.

## ANNEX

Note: Panels report bias-corrected fuzzy regression-discontinuity estimates for either a recorded diabetes diagnosis or a diabetes-medication prescription within six months of the index HbA1c test and outcomes measured during months 12–24. Capped vertical lines are robust 95% confidence intervals. The black diamond is the primary local-linear specification with MSE-selected bandwidth, triangular kernel weights, mass-point adjustment, controls for sex, race, ethnicity, state, index year, age, and age squared, plus the outcome’s baseline value and a baseline-missing indicator when available; standard errors are clustered by care site. Blue circles use 0.75, 1.25, and 1.50 times the primary bandwidth. Gold squares use baseline-only controls or no controls. The green triangle uses a local-quadratic specification. The purple pentagon excludes observations with index HbA1c exactly equal to 6.5%. Gray crosses along the bottom of a panel identify specifications for which the squared robust first-stage z statistic is below 10; their treatment estimates and intervals are suppressed.

**Table A1.** Health and Retirement Study descriptive statistics at the 7.0% cutoff.

| Variable label | Category / statistic | Year 0<br>(1) | Year 2<br>(2) | Year 4<br>(3) | Year 6<br>(4) | Year 8<br>(5) |
| --- | --- | --- | --- | --- | --- | --- |
| **Total observations** | **N** | **386** | **340** | **305** | **266** | **240** |
| Father alive | 0 (No) | 332 | 297 | 272 | 246 | 225 |
|  | 1 (Yes) | 50 | 42 | 32 | 20 | 15 |
| Mother alive | 0 (No) | 271 | 254 | 234 | 211 | 200 |
|  | 1 (Yes) | 111 | 84 | 71 | 55 | 39 |
| Respondent fully retired | 0 (No) | 351 | 295 | 271 | 239 | 213 |
|  | 1 (Yes) | 35 | 45 | 34 | 27 | 27 |
| Respondent partially or fully retired | 0 (No) | 226 | 171 | 143 | 101 | 85 |
|  | 1 (Yes) | 160 | 169 | 162 | 165 | 155 |
| R working for pay | 0 (No) | 199 | 183 | 178 | 176 | 168 |
|  | 1 (Yes) | 187 | 157 | 125 | 90 | 71 |
| R is in the labor force | 0 (No) | 180 | 166 | 162 | 161 | 161 |
|  | 1 (Yes) | 206 | 165 | 137 | 101 | 74 |
| Respondent not working and not retired | 0 (No) | 318 | 299 | 266 | 239 | 213 |
|  | 1 (Yes) | 68 | 41 | 39 | 27 | 27 |
| Respondent not in the LBF and is not retired | 0 (No) | 334 | 306 | 275 | 244 | 215 |
|  | 1 (Yes) | 52 | 34 | 30 | 22 | 25 |
| Health problems limit work | 0 (No) | 262 | 227 | 194 | 156 | 132 |
|  | 1 (Yes) | 91 | 89 | 90 | 94 | 95 |
| R IS RECEIVING SSDI | 0 (No) | 358 | 313 | 278 | 243 | 221 |
|  | 1 (Yes) | 28 | 27 | 27 | 23 | 19 |
| Respondent is married (=1) | 0 (No) | 192 | 168 | 158 | 139 | 134 |
|  | 1 (Yes) | 194 | 172 | 147 | 127 | 106 |
| Respondent takes oral diabetes meds | 0 (No) | 386 | 298 | 245 | 194 | 151 |
|  | 1 (Yes) | 0 | 42 | 60 | 72 | 89 |
| R reports diabetes this wv | 0 (No) | 386 | 257 | 195 | 150 | 117 |
|  | 1 (Yes) | 0 | 83 | 110 | 116 | 123 |
| Genetic data is missing | 0 (No) | 313 | 278 | 251 | 230 | 207 |
|  | 1 (Yes) | 73 | 62 | 54 | 36 | 33 |
| Provider HbA1c value | Mean | 6.758 | — | 6.220 | — | 6.008 |
| Respondent race-specific A1c polygenic score | Mean | 0.124 | 0.148 | 0.110 | 0.089 | 0.052 |
| R Age (years) at lvw BegMon | Mean | 63.764 | 65.629 | 67.115 | 69.271 | 71.812 |
| Hours worked/week main job | Mean | 18.697 | 17.618 | 15.650 | 11.939 | 10.508 |
| log(1 + R_I_EARN) transformation | Mean | 4.689 | 4.085 | 3.906 | 3.255 | 2.794 |
| log(1 + R_I_SSDI) transformation | Mean | 0.662 | 0.923 | 0.873 | 0.816 | 0.754 |
| Any Diff-sum of ADLs /0-5 | Mean | 0.272 | 0.397 | 0.407 | 0.425 | 0.523 |
| Any Diff-sum of IADLs /0-5 | Mean | 0.251 | 0.347 | 0.420 | 0.387 | 0.502 |
| Self-report of health | Mean | 2.948 | 3.000 | 3.030 | 3.019 | 3.196 |
| 27-POINT COGNITION SCORE | Mean | 14.368 | 14.257 | 14.323 | 14.223 | 14.317 |
| # Hospital stays prv 2 yrs (W1: prv 12 mos*) | Mean | 0.330 | 0.386 | 0.411 | 0.534 | 0.710 |

*Note:* Baseline HbA1c window: 6.5–7.5%. Binary variables have separate frequency rows for 0 (No) and 1 (Yes); continuous variables report means. N denotes total observations. An em dash preserves a blank or period entry in the source. Counts can sum to less than total observations because of missing values. Year 0 is baseline; subsequent columns are years after the initial test.

**Table A2.** Health and Retirement Study descriptive statistics at the 6.5% cutoff.

| Variable label | Category /<br>statistic | Year 0<br>(1) | Year 2<br>(2) | Year 4<br>(3) | Year 6<br>(4) | Year 8<br>(5) |
| --- | --- | --- | --- | --- | --- | --- |
| **Total observations** | N | 1,921 | 1,727 | 1,554 | 1,332 | 1,162 |
| Father alive | 0 (No) | 1,667 | 1,523 | 1,402 | 1,238 | 1,100 |
|  | 1 (Yes) | 233 | 193 | 141 | 89 | 58 |
| Mother alive | 0 (No) | 1,351 | 1,268 | 1,183 | 1,062 | 976 |
|  | 1 (Yes) | 550 | 442 | 352 | 260 | 177 |
| Respondent fully retired | 0 (No) | 1,710 | 1,524 | 1,366 | 1,165 | 1,010 |
|  | 1 (Yes) | 211 | 203 | 188 | 167 | 152 |
| Respondent partially or fully retired | 0 (No) | 1,058 | 843 | 660 | 461 | 334 |
|  | 1 (Yes) | 863 | 884 | 894 | 871 | 828 |
| R working for pay | 0 (No) | 990 | 955 | 920 | 873 | 810 |
|  | 1 (Yes) | 931 | 772 | 627 | 457 | 348 |
| R is in the labor force | 0 (No) | 911 | 878 | 842 | 791 | 762 |
|  | 1 (Yes) | 1,010 | 827 | 674 | 496 | 364 |
| Respondent not working and not retired | 0 (No) | 1,628 | 1,519 | 1,404 | 1,222 | 1,074 |
|  | 1 (Yes) | 293 | 208 | 150 | 110 | 88 |
| Respondent not in the LBF and is not retired | 0 (No) | 1,689 | 1,555 | 1,434 | 1,247 | 1,084 |
|  | 1 (Yes) | 232 | 172 | 120 | 85 | 78 |
| Health problems limit work | 0 (No) | 1,316 | 1,125 | 976 | 783 | 638 |
|  | 1 (Yes) | 456 | 489 | 481 | 476 | 468 |
| R IS RECEIVING SSDI | 0 (No) | 1,816 | 1,621 | 1,457 | 1,249 | 1,092 |
|  | 1 (Yes) | 105 | 106 | 97 | 83 | 70 |
| Respondent is married | 0 (No) | 840 | 794 | 763 | 664 | 613 |
|  | 1 (Yes) | 1,081 | 933 | 791 | 668 | 549 |
| Respondent takes oral diabetes meds | 0 (No) | 1,921 | 1,640 | 1,437 | 1,180 | 951 |
|  | 1 (Yes) | — | 87 | 117 | 152 | 211 |
| R reports diabetes this wv | 0 (No) | 1,921 | 1,559 | 1,290 | 1,038 | 847 |
|  | 1 (Yes) | — | 168 | 262 | 294 | 313 |
| Genetic data is missing | 0 (No) | 1,580 | 1,427 | 1,281 | 1,139 | 1,014 |
|  | 1 (Yes) | 341 | 300 | 273 | 193 | 148 |
| Provider HbA1c value | Mean | 6.231 | — | 5.866 | — | 5.702 |
| Respondent race-specific A1c polygenic score | Mean | 0.084 | 0.086 | 0.062 | 0.058 | 0.065 |
| R Age (years) at lwv BegMon | Mean | 64.341 | 66.252 | 68.028 | 70.158 | 72.065 |
| Hours worked/week main job | Mean | 17.701 | 16.318 | 14.356 | 11.898 | 10.007 |
| log(1 + R_I_EARN) transformation | Mean | 4.602 | 4.084 | 3.847 | 3.218 | 2.884 |
| log(1 + R_I_SSDI) transformation | Mean | 0.581 | 0.653 | 0.693 | 0.700 | 0.716 |
| Any Diff-sum of ADLs /0-5 | Mean | 0.250 | 0.321 | 0.373 | 0.405 | 0.446 |
| Any Diff-sum of IADLs /0-5 | Mean | 0.214 | 0.298 | 0.387 | 0.403 | 0.445 |
| Self-report of health | Mean | 2.799 | 2.859 | 2.898 | 2.895 | 2.997 |
| 27-POINT COGNITION SCORE | Mean | 14.675 | 14.827 | 14.436 | 14.539 | 14.323 |
| # Hospital stays prv 2 yrs (W1: prv 12 mos*) | Mean | 0.362 | 0.445 | 0.402 | 0.444 | 0.536 |

*Note:* Baseline HbA1c window: 6.0–7.0%. Binary variables have separate frequency rows for 0 (No) and 1 (Yes); continuous variables report means. N denotes total observations. An em dash preserves a blank or period entry in the source. Counts can sum to less than total observations because of missing values. Year 0 is baseline; subsequent columns are years after the initial test.

**Fig. A1.**
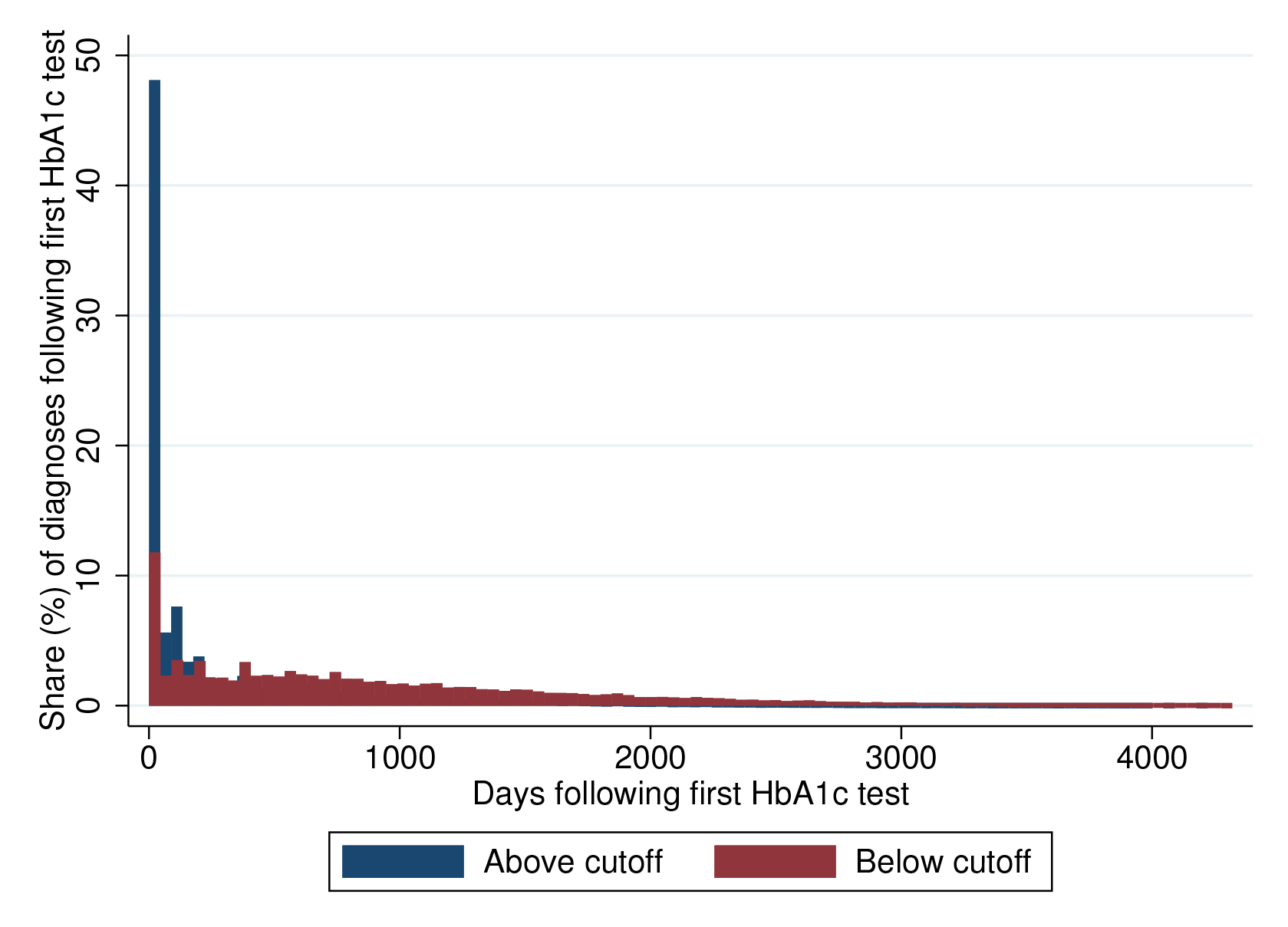
American Family Cohort: Distribution of diabetes diagnoses in the time following first HbA1c test. **Constant-follow-up analytic cohort**. For the persistence figures below, the AFC analytic cohort is restricted to patients observed for at least 96 months after the index HbA1c test. Requiring observation through the longest reported horizon keeps the set of patients constant across the 30-day, 90-day, and 6-through 96-month estimates, so changes over time are not driven by progressively selective loss of observable patients.

**Fig. A2.**
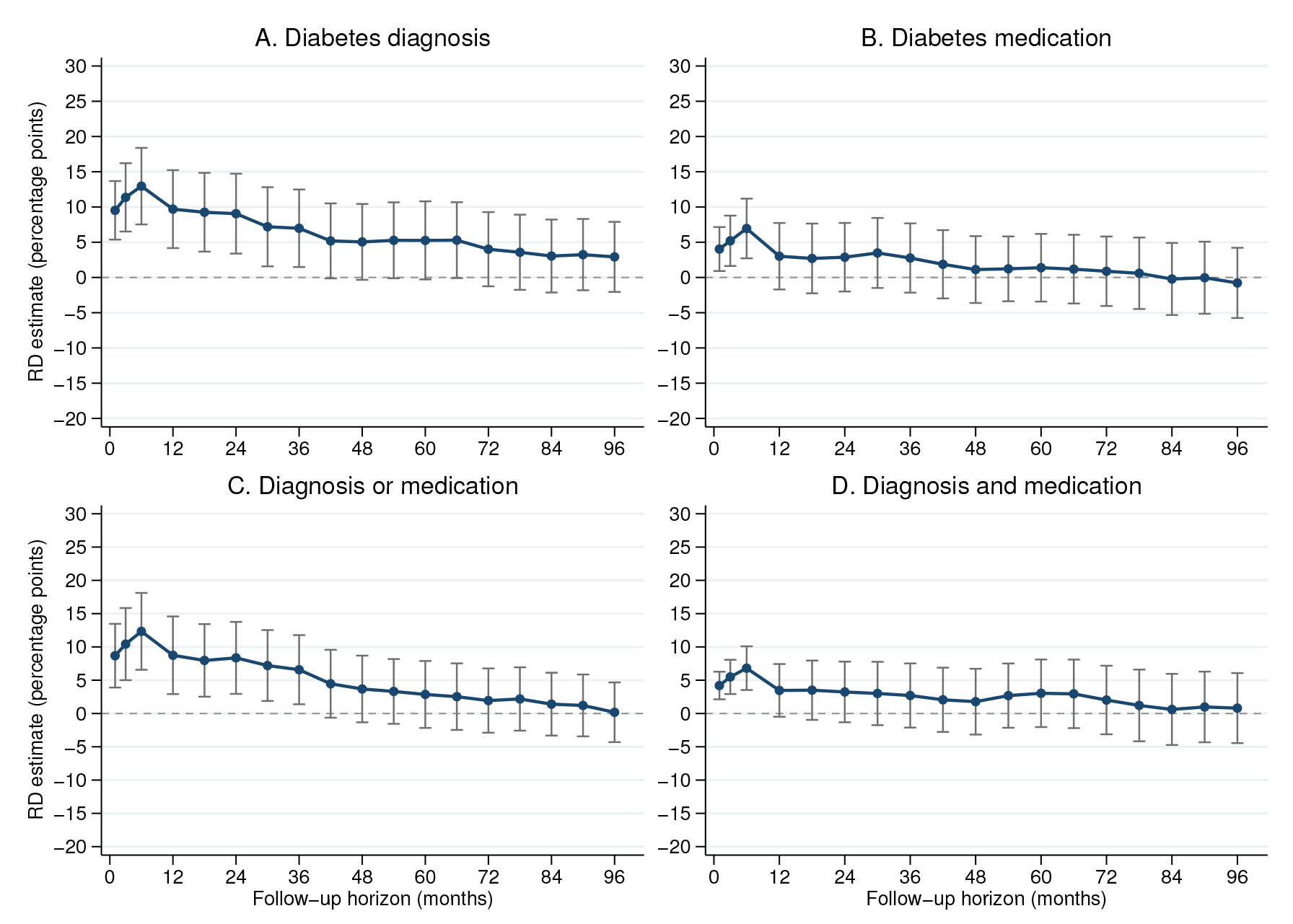
Persistence of diagnosis and medication discontinuities at the HbA1c threshold: Full AFC sample. *Note*: Each point is a robust bias-corrected local-linear regression-discontinuity estimate of the change, in percentage points, in the cumulative probability of the indicated endpoint at the HbA1c diagnostic threshold of 6.5%; capped vertical lines show robust 95% confidence intervals. The four panels distinguish a recorded diabetes diagnosis, a diabetes-medication record, either event, and both events. Positive estimates indicate a higher cumulative probability immediately above the diagnostic threshold. The analysis uses the full AFC analytic cohort of 90,214 patients meeting the constant-follow-up definition given above. Outcomes are measured by 30 days, 90 days, and at six-month intervals from 6 through 96 months. All specifications use a local-linear polynomial with a local-quadratic bias correction, a triangular kernel, adjustment for mass points in the running variable, and standard errors clustered by care site. For each endpoint, the MSE-optimal estimation bandwidth and its associated bias bandwidth were selected using the six-month outcome and then held fixed across all follow-up horizons, so changes in the plotted estimates do not reflect changes in bandwidth. No additional covariates are included. The y-axis runs from −20 to 30 percentage points, with labels every 5 percentage points, in every panel of every persistence figure; confidence intervals extending outside this display range are truncated at the corresponding boundary.

**Fig. A3.**
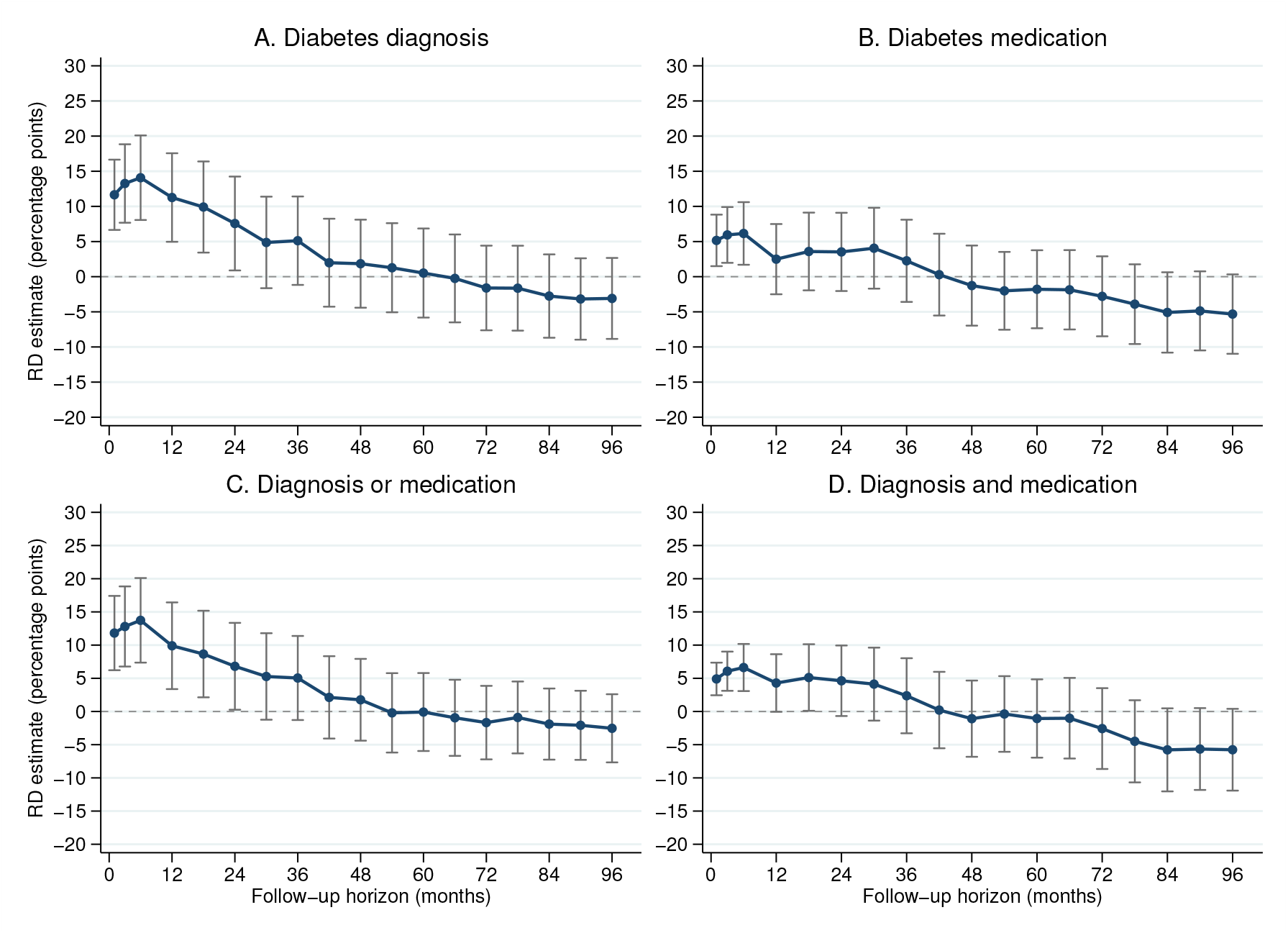
Persistence of diagnosis and medication discontinuities at the HbA1c threshold: Men. *Note*: Each point is a robust bias-corrected local-linear regression-discontinuity estimate of the change, in percentage points, in the cumulative probability of the indicated endpoint at the HbA1c diagnostic threshold of 6.5%; capped vertical lines show robust 95% confidence intervals. The four panels distinguish a recorded diabetes diagnosis, a diabetes-medication record, either event, and both events. Positive estimates indicate a higher cumulative probability immediately above the diagnostic threshold. The analysis is restricted to men within the AFC analytic cohort defined above and contains 40,318 patients. Outcomes are measured by 30 days, 90 days, and at six-month intervals from 6 through 96 months. All specifications use a local-linear polynomial with a local-quadratic bias correction, a triangular kernel, adjustment for mass points in the running variable, and standard errors clustered by care site. For each endpoint, the MSE-optimal estimation bandwidth and its associated bias bandwidth were selected using the six-month outcome and then held fixed across all follow-up horizons. No additional covariates are included. The y-axis runs from −20 to 30 percentage points, with labels every 5 percentage points, in every panel of every persistence figure; confidence intervals extending outside this display range are truncated at the corresponding boundary. Survey names in the age-restricted captions identify comparator age ranges; all estimates shown here use AFC data.

**Fig. A4.**
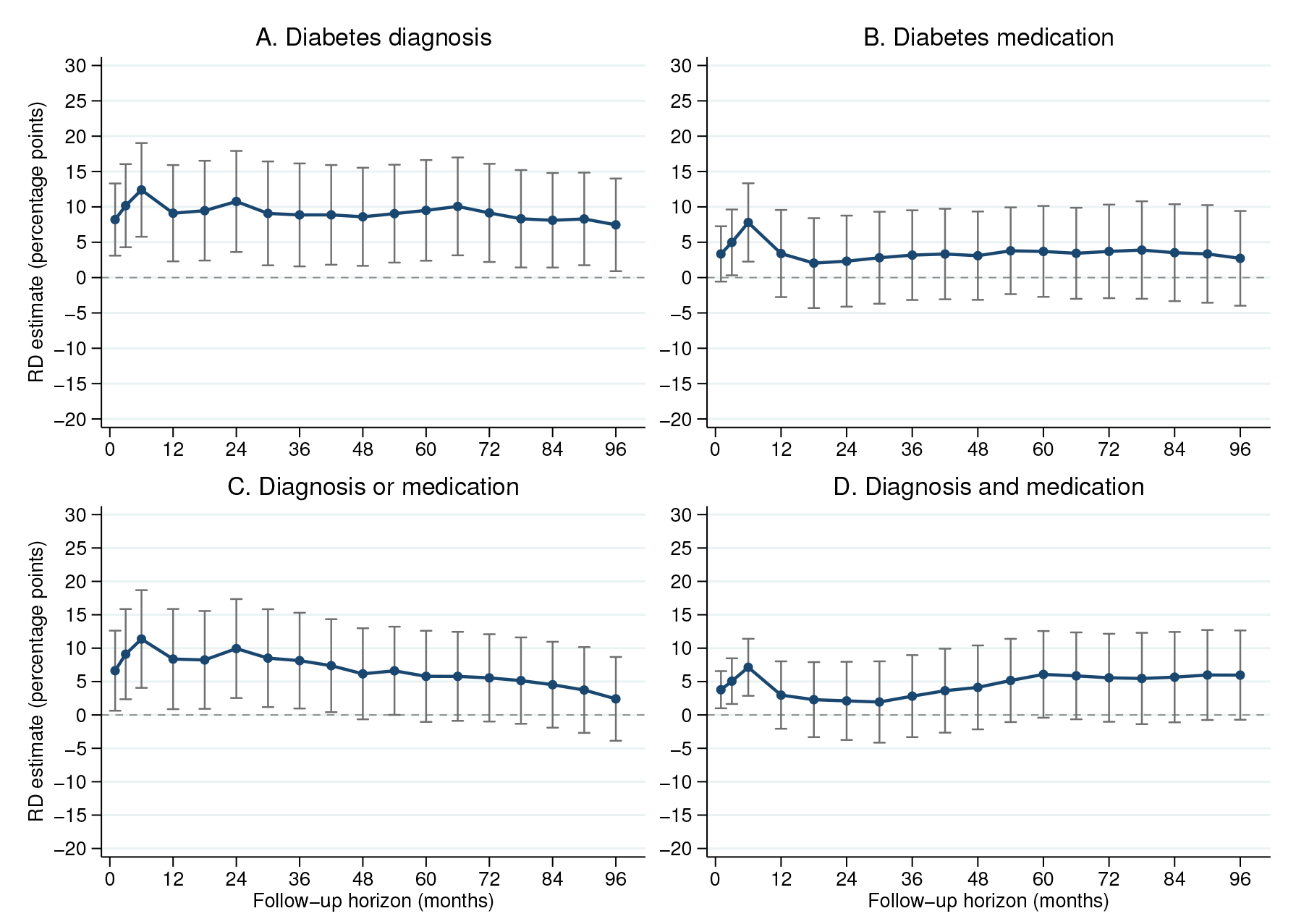
Persistence of diagnosis and medication discontinuities at the HbA1c threshold: Women. *Note*: Each point is a robust bias-corrected local-linear regression-discontinuity estimate of the change, in percentage points, in the cumulative probability of the indicated endpoint at the HbA1c diagnostic threshold of 6.5%; capped vertical lines show robust 95% confidence intervals. The four panels distinguish a recorded diabetes diagnosis, a diabetes-medication record, either event, and both events. Positive estimates indicate a higher cumulative probability immediately above the diagnostic threshold. The analysis is restricted to women within the AFC analytic cohort defined above and contains 49,889 patients. Outcomes are measured by 30 days, 90 days, and at six-month intervals from 6 through 96 months. All specifications use a local-linear polynomial with a local-quadratic bias correction, a triangular kernel, adjustment for mass points in the running variable, and standard errors clustered by care site. For each endpoint, the MSE-optimal estimation bandwidth and its associated bias bandwidth were selected using the six-month outcome and then held fixed across all follow-up horizons. No additional covariates are included. The y-axis runs from −20 to 30 percentage points, with labels every 5 percentage points, in every panel of every persistence figure; confidence intervals extending outside this display range are truncated at the corresponding boundary. Survey names in the age-restricted captions identify comparator age ranges; all estimates shown here use AFC data.

**Fig. A5.**
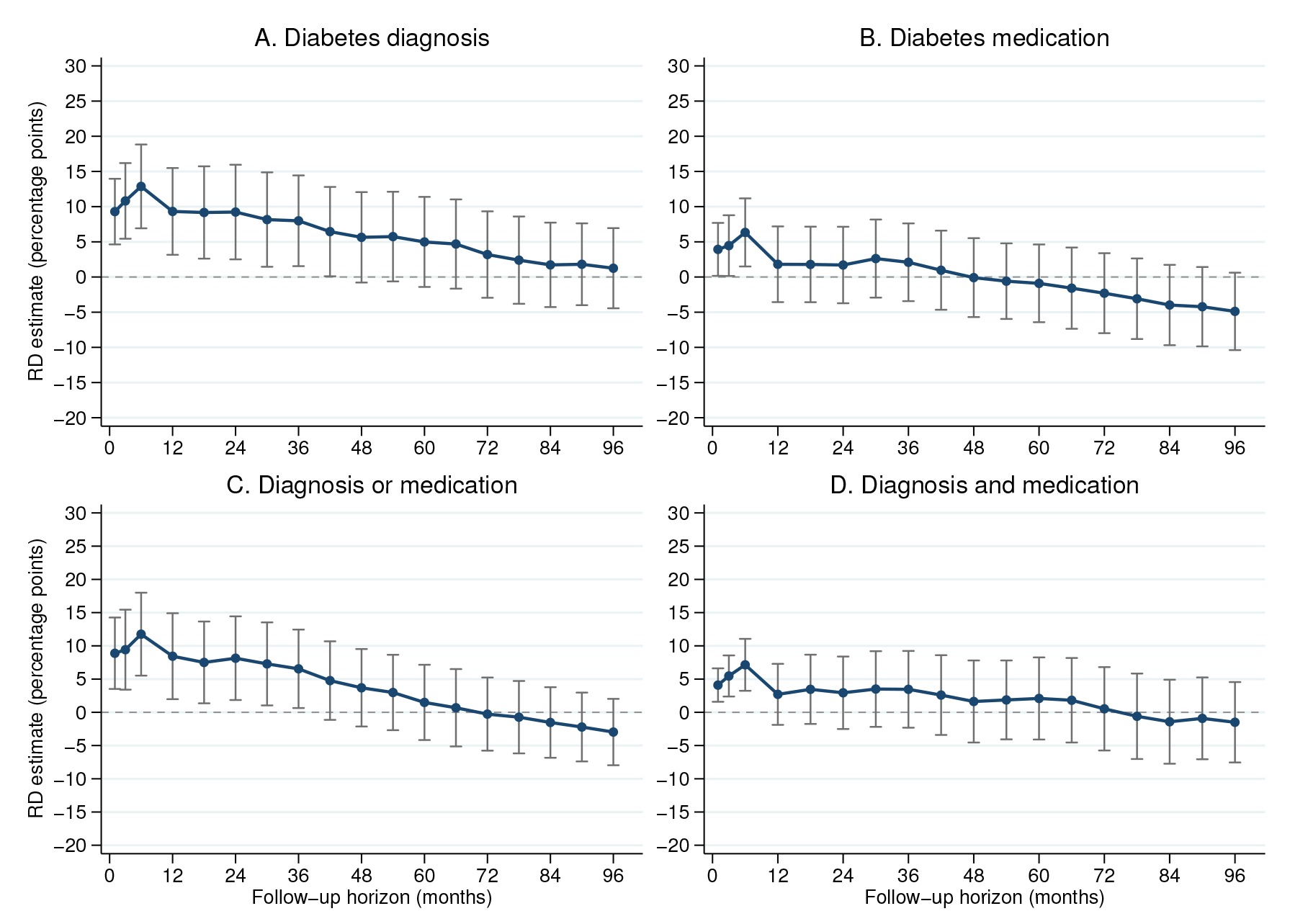
Persistence of diagnosis and medication discontinuities at the HbA1c threshold: White. *Note*: Each point is a robust bias-corrected local-linear regression-discontinuity estimate of the change, in percentage points, in the cumulative probability of the indicated endpoint at the HbA1c diagnostic threshold of 6.5%; capped vertical lines show robust 95% confidence intervals. The four panels distinguish a recorded diabetes diagnosis, a diabetes-medication record, either event, and both events. Positive estimates indicate a higher cumulative probability immediately above the diagnostic threshold. The analysis is restricted to patients coded as White within the AFC analytic cohort defined above and contains 72,810 patients. Outcomes are measured by 30 days, 90 days, and at six-month intervals from 6 through 96 months. All specifications use a local-linear polynomial with a local-quadratic bias correction, a triangular kernel, adjustment for mass points in the running variable, and standard errors clustered by care site. For each endpoint, the MSE-optimal estimation bandwidth and its associated bias bandwidth were selected using the six-month outcome and then held fixed across all follow-up horizons. No additional covariates are included. The y-axis runs from −20 to 30 percentage points, with labels every 5 percentage points, in every panel of every persistence figure; confidence intervals extending outside this display range are truncated at the corresponding boundary. Survey names in the age-restricted captions identify comparator age ranges; all estimates shown here use AFC data.

**Fig. A6.**
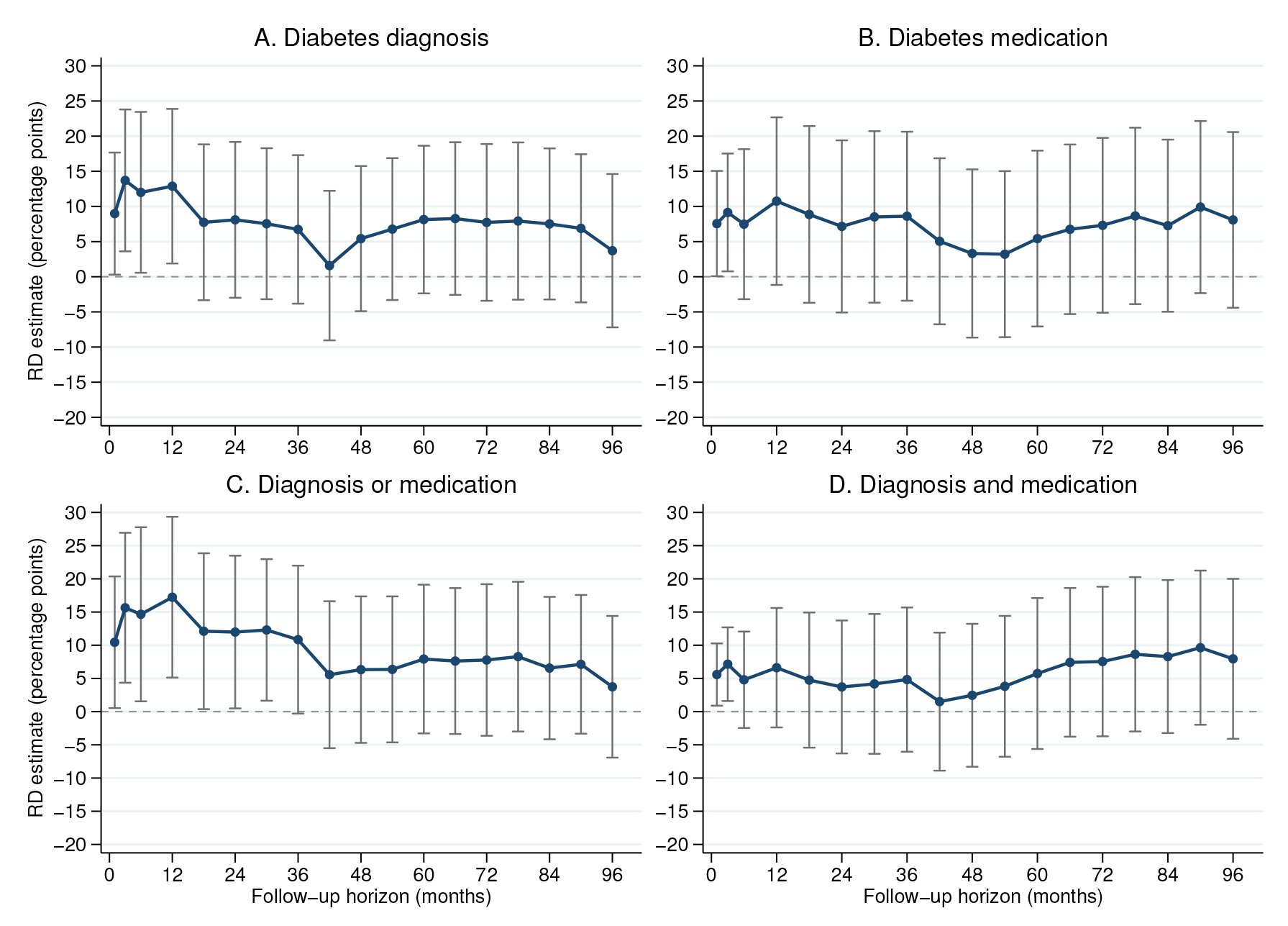
Persistence of diagnosis and medication discontinuities at the HbA1c threshold: Black. *Note*: Each point is a robust bias-corrected local-linear regression-discontinuity estimate of the change, in percentage points, in the cumulative probability of the indicated endpoint at the HbA1c diagnostic threshold of 6.5%; capped vertical lines show robust 95% confidence intervals. The four panels distinguish a recorded diabetes diagnosis, a diabetes-medication record, either event, and both events. Positive estimates indicate a higher cumulative probability immediately above the diagnostic threshold. The analysis is restricted to patients coded as Black within the AFC analytic cohort defined above and contains 6,465 patients. Outcomes are measured by 30 days, 90 days, and at six-month intervals from 6 through 96 months. All specifications use a local-linear polynomial with a local-quadratic bias correction, a triangular kernel, adjustment for mass points in the running variable, and standard errors clustered by care site. For each endpoint, the MSE-optimal estimation bandwidth and its associated bias bandwidth were selected using the six-month outcome and then held fixed across all follow-up horizons. No additional covariates are included. The y-axis runs from −20 to 30 percentage points, with labels every 5 percentage points, in every panel of every persistence figure; confidence intervals extending outside this display range are truncated at the corresponding boundary. Survey names in the age-restricted captions identify comparator age ranges; all estimates shown here use AFC data.

**Fig. A7.**
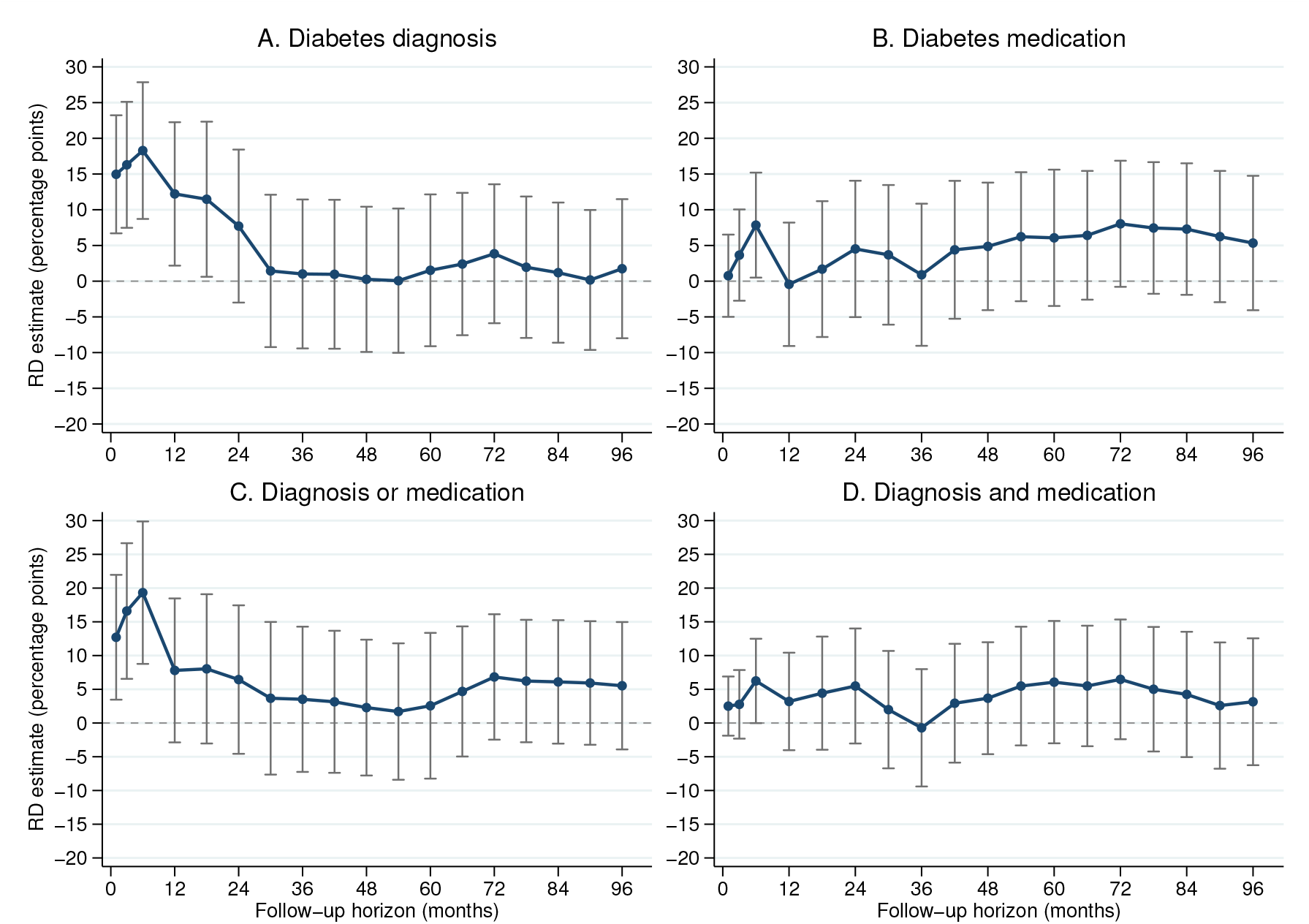
Persistence of diagnosis and medication discontinuities at the HbA1c threshold: Neither White nor Black. *Note*: Each point is a robust bias-corrected local-linear regression-discontinuity estimate of the change, in percentage points, in the cumulative probability of the indicated endpoint at the HbA1c diagnostic threshold of 6.5%; capped vertical lines show robust 95% confidence intervals. The four panels distinguish a recorded diabetes diagnosis, a diabetes-medication record, either event, and both events. Positive estimates indicate a higher cumulative probability immediately above the diagnostic threshold. The analysis is restricted to patients with observed race who were coded as neither White nor Black within the AFC analytic cohort defined above and contains 9,503 patients. Outcomes are measured by 30 days, 90 days, and at six-month intervals from 6 through 96 months. All specifications use a local-linear polynomial with a local-quadratic bias correction, a triangular kernel, adjustment for mass points in the running variable, and standard errors clustered by care site. For each endpoint, the MSE-optimal estimation bandwidth and its associated bias bandwidth were selected using the six-month outcome and then held fixed across all follow-up horizons. No additional covariates are included. The y-axis runs from −20 to 30 percentage points, with labels every 5 percentage points, in every panel of every persistence figure; confidence intervals extending outside this display range are truncated at the corresponding boundary. Survey names in the age-restricted captions identify comparator age ranges; all estimates shown here use AFC data.

**Fig. A8.**
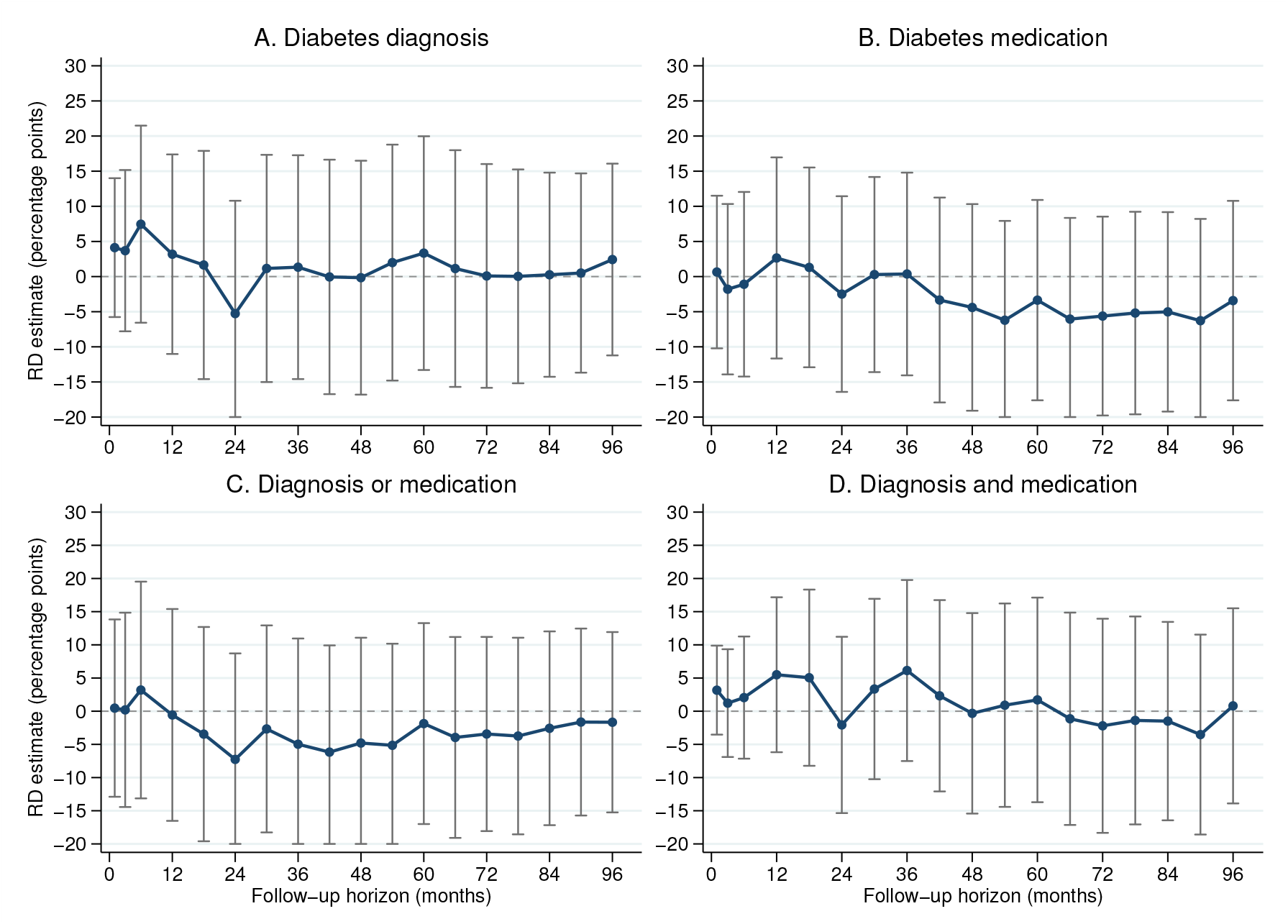
Persistence of diagnosis and medication discontinuities at the HbA1c threshold: Age 18–40. *Note*: Each point is a robust bias-corrected local-linear regression-discontinuity estimate of the change, in percentage points, in the cumulative probability of the indicated endpoint at the HbA1c diagnostic threshold of 6.5%; capped vertical lines show robust 95% confidence intervals. The four panels distinguish a recorded diabetes diagnosis, a diabetes-medication record, either event, and both events. Positive estimates indicate a higher cumulative probability immediately above the diagnostic threshold. The analysis is restricted to patients aged 18–40 at the index test within the AFC analytic cohort defined above and contains 11,606 patients. Outcomes are measured by 30 days, 90 days, and at six-month intervals from 6 through 96 months. All specifications use a local-linear polynomial with a local-quadratic bias correction, a triangular kernel, adjustment for mass points in the running variable, and standard errors clustered by care site. For each endpoint, the MSE-optimal estimation bandwidth and its associated bias bandwidth were selected using the six-month outcome and then held fixed across all follow-up horizons. No additional covariates are included. The y-axis runs from −20 to 30 percentage points, with labels every 5 percentage points, in every panel of every persistence figure; confidence intervals extending outside this display range are truncated at the corresponding boundary. Survey names in the age-restricted captions identify comparator age ranges; all estimates shown here use AFC data.

**Fig. A9.**
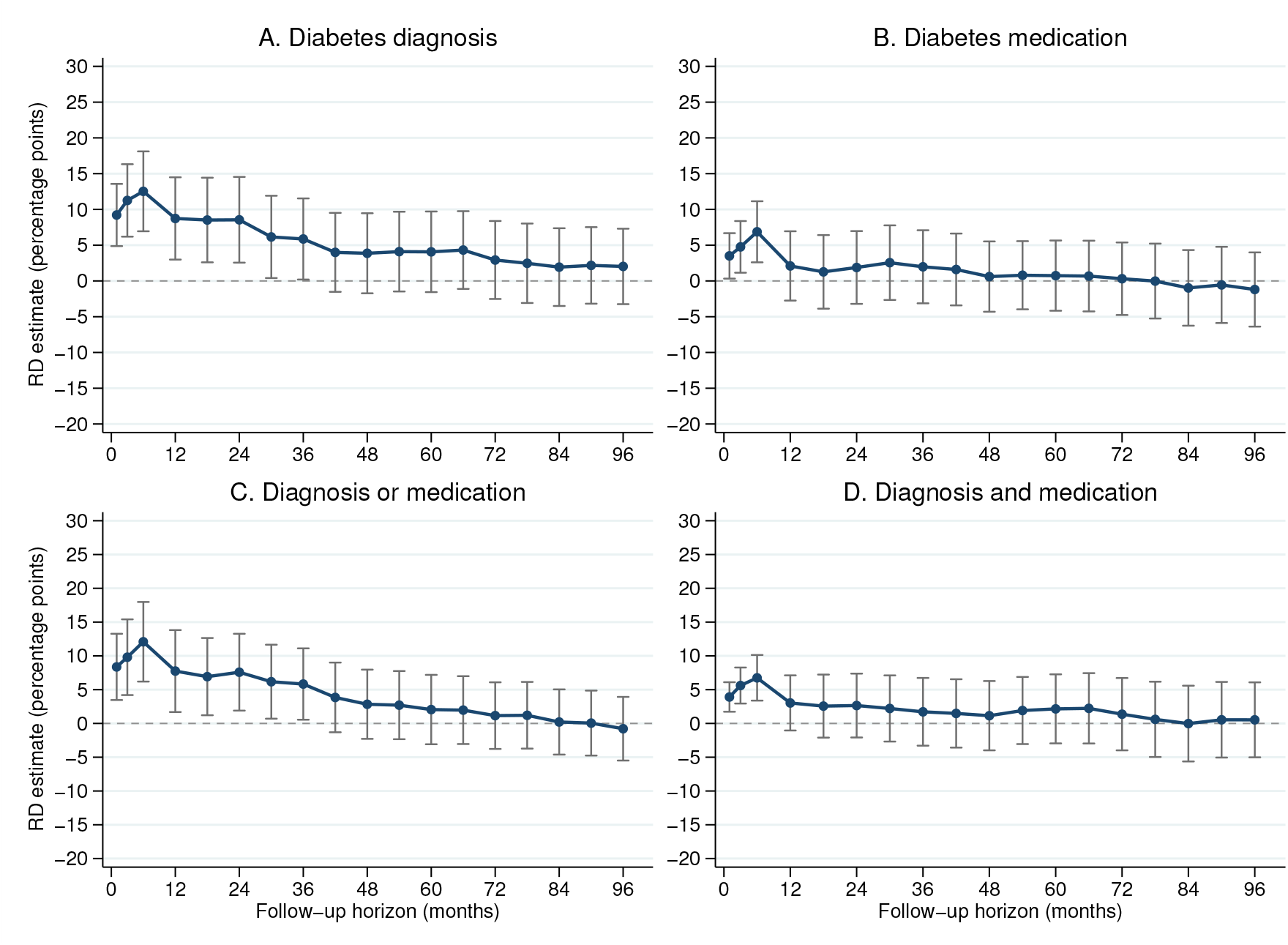
Persistence of diagnosis and medication discontinuities at the HbA1c threshold: Age 50 or older (HRS-comparable age range) *Note*: Each point is a robust bias-corrected local-linear regression-discontinuity estimate of the change, in percentage points, in the cumulative probability of the indicated endpoint at the HbA1c diagnostic threshold of 6.5%; capped vertical lines show robust 95% confidence intervals. The four panels distinguish a recorded diabetes diagnosis, a diabetes-medication record, either event, and both events. Positive estimates indicate a higher cumulative probability immediately above the diagnostic threshold. The analysis is restricted to patients aged 50–120 at the index test within the AFC analytic cohort defined above and contains 62,801 patients. Outcomes are measured by 30 days, 90 days, and at six-month intervals from 6 through 96 months. All specifications use a local-linear polynomial with a local-quadratic bias correction, a triangular kernel, adjustment for mass points in the running variable, and standard errors clustered by care site. For each endpoint, the MSE-optimal estimation bandwidth and its associated bias bandwidth were selected using the six-month outcome and then held fixed across all follow-up horizons. No additional covariates are included. The y-axis runs from −20 to 30 percentage points, with labels every 5 percentage points, in every panel of every persistence figure; confidence intervals extending outside this display range are truncated at the corresponding boundary. Survey names in the age-restricted captions identify comparator age ranges; all estimates shown here use AFC data.

**Table A3.**
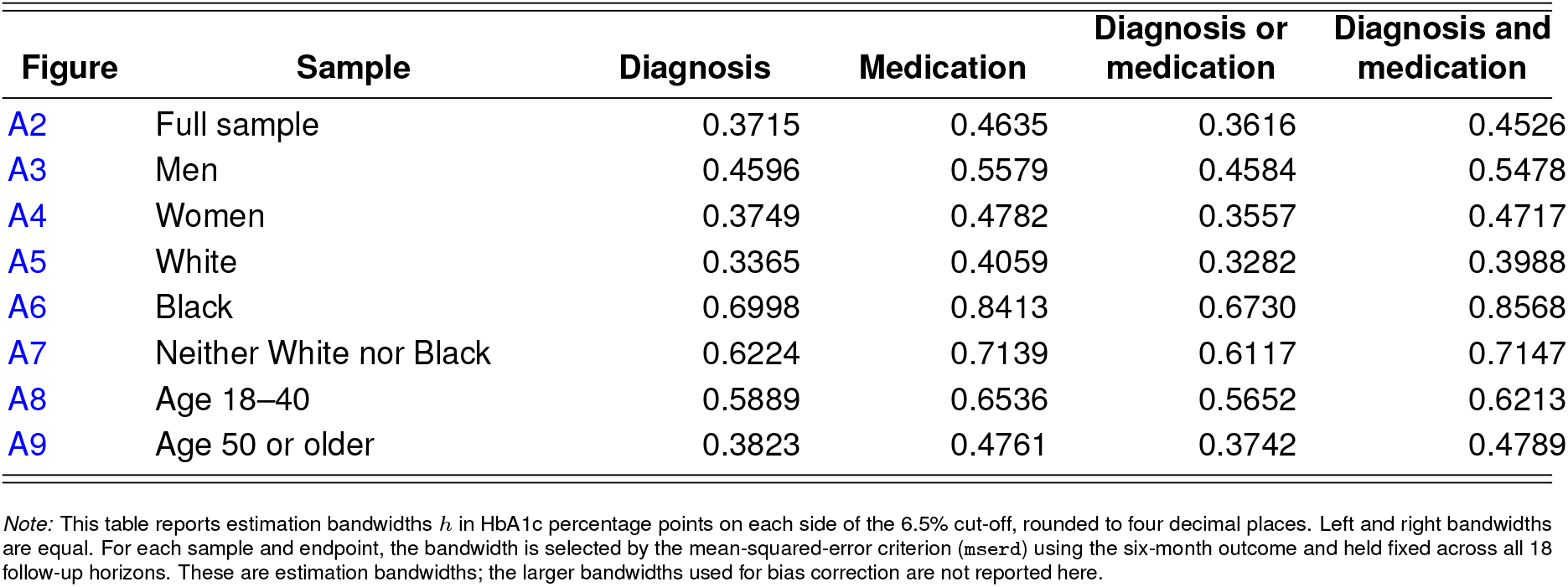
Selected estimation bandwidths around the 6.5 percent threshold for AFC persistence figures.

**Table A4.** Robust first-stage diagnostics for dynamic AFC estimates.

Squared robust first-stage $z$ statistic ( $z^2$ )
| Outcome | Outcome window | Diagnosis |  |  |  | Prescription |  |  |  | Either diagnosis or prescription |  |  |  | Both diagnosis and prescription |  |  |  |
| --- | --- | --- | --- | --- | --- | --- | --- | --- | --- | --- | --- | --- | --- | --- | --- | --- | --- |
|  |  | 6 m | 12 m | 24 m | 36 m | 6 m | 12 m | 24 m | 36 m | 6 m | 12 m | 24 m | 36 m | 6 m | 12 m | 24 m | 36 m |
|  |  | (1) | (2) | (3) | (4) | (5) | (6) | (7) | (8) | (9) | (10) | (11) | (12) | (13) | (14) | (15) | (16) |
| HbA1c | 6–12 | 30.9 | – | – | – | 0.0 | – | – | – | 15.6 | – | – | – | 2.8 | – | – | – |
| HbA1c | 12–24 | 67.8 | 40.5 | – | – | 0.2 | 0.0 | – | – | 40.0 | 27.1 | – | – | 7.2 | 2.0 | – | – |
| HbA1c | 24–36 | 59.2 | 34.3 | 13.1 | – | 1.0 | 0.3 | 0.1 | – | 35.7 | 23.5 | 8.1 | – | 8.5 | 2.6 | 0.7 | – |
| HbA1c | 36–48 | 48.5 | 28.6 | 9.9 | 4.7 | 3.0 | 1.0 | 0.0 | 0.0 | 34.2 | 23.1 | 7.2 | 5.3 | 10.3 | 1.6 | 0.9 | 0.0 |
| HbA1c | 60–72 | 28.3 | 25.5 | 13.5 | 8.1 | 1.6 | 0.9 | 0.1 | 0.2 | 27.8 | 19.2 | 10.2 | 7.2 | 5.5 | 2.8 | 0.9 | 0.3 |
| HbA1c | 84–96 | 20.3 | 15.3 | 13.2 | 10.1 | 6.6 | 1.2 | 0.6 | 0.9 | 14.5 | 13.4 | 11.0 | 10.8 | 11.7 | 2.4 | 2.3 | 1.5 |
| Glucose | 6–12 | 30.9 | – | – | – | 0.9 | – | – | – | 19.7 | – | – | – | 5.1 | – | – | – |
| Glucose | 12–24 | 30.2 | 25.6 | – | – | 1.1 | 0.2 | – | – | 17.7 | 11.1 | – | – | 6.7 | 0.0 | – | – |
| Glucose | 24–36 | 34.8 | 18.2 | 4.3 | – | 0.0 | 0.0 | 0.4 | – | 21.2 | 8.6 | 1.1 | – | 3.7 | 0.8 | 0.2 | – |
| Glucose | 36–48 | 22.8 | 11.2 | 3.2 | 1.1 | 2.3 | 0.1 | 0.7 | 1.2 | 16.3 | 9.2 | 2.1 | 1.4 | 0.8 | 0.0 | 2.9 | 0.0 |
| Glucose | 60–72 | 31.7 | 14.7 | 8.9 | 6.8 | 1.9 | 0.5 | 0.1 | 0.4 | 22.0 | 11.4 | 7.5 | 7.3 | 12.2 | 7.5 | 0.8 | 0.4 |
| Glucose | 84–96 | 20.3 | 16.3 | 6.4 | 6.8 | 13.6 | 0.9 | 0.6 | 1.9 | 32.1 | 11.9 | 7.4 | 7.7 | 15.7 | 2.1 | 1.0 | 0.8 |
| BMI | 6–12 | 80.1 | – | – | – | 2.7 | – | – | – | 59.8 | – | – | – | 16.4 | – | – | – |
| BMI | 12–24 | 95.6 | 59.7 | – | – | 1.7 | 0.0 | – | – | 57.9 | 56.2 | – | – | 15.4 | 6.2 | – | – |
| BMI | 24–36 | 98.7 | 59.8 | 25.2 | – | 3.2 | 0.6 | 0.4 | – | 88.1 | 33.8 | 8.4 | – | 15.0 | 6.2 | 1.2 | – |
| BMI | 36–48 | 75.0 | 44.7 | 18.3 | 7.3 | 2.2 | 0.4 | 0.8 | 0.2 | 50.3 | 31.4 | 13.4 | 7.7 | 20.4 | 3.2 | 2.1 | 0.0 |
| BMI | 60–72 | 40.5 | 33.6 | 11.5 | 3.3 | 1.7 | 0.2 | 0.0 | 0.0 | 26.0 | 23.0 | 5.3 | 2.4 | 8.9 | 2.4 | 3.3 | 0.1 |
| BMI | 84–96 | 29.4 | 12.5 | 10.2 | 2.1 | 2.3 | 1.1 | 0.1 | 0.1 | 21.6 | 13.8 | 2.0 | 0.4 | 18.5 | 5.6 | 2.5 | 0.6 |
| Weight | 6–12 | 80.1 | – | – | – | 2.2 | – | – | – | 67.5 | – | – | – | 20.6 | – | – | – |
| Weight | 12–24 | 92.6 | 60.6 | – | – | 2.7 | 0.0 | – | – | 53.4 | 51.9 | – | – | 22.2 | 4.1 | – | – |
| Weight | 24–36 | 120.7 | 61.4 | 28.5 | – | 2.5 | 0.0 | 0.2 | – | 77.3 | 47.2 | 11.6 | – | 15.0 | 13.1 | 3.0 | – |
| Weight | 36–48 | 68.6 | 41.3 | 18.4 | 9.7 | 4.1 | 1.6 | 1.0 | 0.6 | 72.1 | 34.9 | 20.9 | 11.4 | 7.7 | 2.3 | 2.8 | 0.1 |
| Weight | 60–72 | 45.6 | 33.7 | 12.4 | 7.5 | 0.9 | 0.7 | 0.1 | 0.1 | 31.0 | 25.7 | 10.5 | 8.4 | 7.0 | 2.6 | 0.6 | 0.1 |
| Weight | 84–96 | 33.7 | 22.3 | 24.8 | 6.3 | 9.9 | 1.0 | 0.0 | 0.4 | 25.9 | 17.1 | 8.4 | 6.2 | 16.6 | 3.7 | 1.6 | 0.6 |
| Systolic blood pressure | 6–12 | 81.6 | – | – | – | 2.0 | – | – | – | 75.7 | – | – | – | 21.9 | – | – | – |
| Systolic blood pressure | 12–24 | 97.4 | 88.2 | – | – | 2.4 | 0.0 | – | – | 80.0 | 48.6 | – | – | 26.0 | 3.7 | – | – |
| Systolic blood pressure | 24–36 | 136.8 | 81.8 | 38.7 | – | 7.7 | 0.9 | 0.0 | – | 102.1 | 51.9 | 12.5 | – | 25.1 | 4.1 | 3.8 | – |
| Systolic blood pressure | 36–48 | 72.0 | 41.9 | 19.7 | 9.9 | 5.5 | 1.3 | 0.8 | 0.2 | 76.4 | 39.3 | 18.5 | 11.3 | 15.7 | 2.9 | 0.9 | 0.1 |
| Systolic blood pressure | 60–72 | 48.5 | 33.8 | 11.8 | 7.3 | 2.5 | 0.9 | 0.0 | 0.0 | 44.8 | 25.4 | 9.1 | 7.8 | 7.4 | 2.3 | 0.7 | 0.1 |
| Systolic blood pressure | 84–96 | 29.8 | 19.7 | 11.3 | 4.6 | 7.2 | 0.0 | 0.0 | 0.2 | 21.5 | 12.4 | 7.0 | 5.1 | 7.6 | 2.0 | 0.9 | 0.2 |
| Diastolic blood pressure | 6–12 | 106.7 | – | – | – | 1.8 | – | – | – | 69.4 | – | – | – | 21.4 | – | – | – |
| Diastolic blood pressure | 12–24 | 92.7 | 55.4 | – | – | 2.2 | 0.1 | – | – | 51.8 | 32.9 | – | – | 22.3 | 3.5 | – | – |
| Diastolic blood pressure | 24–36 | 136.2 | 83.8 | 41.0 | – | 3.5 | 0.0 | 0.1 | – | 88.2 | 44.4 | 12.1 | – | 26.3 | 4.1 | 4.8 | – |
| Diastolic blood pressure | 36–48 | 72.3 | 40.9 | 26.1 | 9.6 | 6.4 | 2.2 | 1.4 | 0.2 | 79.3 | 41.3 | 19.9 | 10.8 | 20.7 | 2.9 | 2.4 | 0.1 |
| Diastolic blood pressure | 60–72 | 47.7 | 30.7 | 11.1 | 7.2 | 2.5 | 0.3 | 0.0 | 0.0 | 36.9 | 22.0 | 8.8 | 7.8 | 7.2 | 2.3 | 0.5 | 0.1 |
| Diastolic blood pressure | 84–96 | 31.0 | 17.8 | 10.9 | 4.5 | 8.0 | 0.1 | 0.0 | 0.3 | 20.9 | 12.4 | 7.2 | 5.0 | 14.3 | 3.8 | 0.8 | 0.3 |
| LDL cholesterol | 6–12 | 30.1 | – | – | – | 2.0 | – | – | – | 14.0 | – | – | – | 10.0 | – | – | – |
| LDL cholesterol | 12–24 | 54.0 | 35.3 | – | – | 0.9 | 0.5 | – | – | 26.5 | 23.6 | – | – | 4.6 | 2.0 | – | – |
| LDL cholesterol | 24–36 | 48.0 | 25.8 | 9.5 | – | 0.0 | 0.0 | 0.7 | – | 20.4 | 12.7 | 3.1 | – | 4.1 | 2.2 | 0.4 | – |
| LDL cholesterol | 36–48 | 25.2 | 16.0 | 7.0 | 7.5 | 0.0 | 0.0 | 0.4 | 0.1 | 16.5 | 13.5 | 5.7 | 6.7 | 0.6 | 0.2 | 0.1 | 0.0 |
| LDL cholesterol | 60–72 | 33.2 | 25.7 | 13.6 | 11.7 | 4.1 | 4.1 | 2.4 | 2.6 | 32.7 | 24.5 | 19.0 | 15.4 | 6.6 | 5.1 | 3.3 | 1.8 |
| LDL cholesterol | 84–96 | 20.1 | 16.4 | 8.8 | 7.8 | 8.8 | 1.8 | 1.0 | 1.3 | 25.9 | 16.8 | 8.7 | 6.8 | 8.3 | 2.4 | 1.9 | 2.8 |
| HbA1c tests | 6–12 | 140.7 | – | – | – | 20.0 | – | – | – | 162.9 | – | – | – | 44.4 | – | – | – |
| HbA1c tests | 12–24 | 106.9 | 55.9 | – | – | 23.7 | 1.8 | – | – | 95.4 | 41.1 | – | – | 41.7 | 6.1 | – | – |
| HbA1c tests | 24–36 | 99.3 | 54.8 | 21.2 | – | 20.8 | 1.9 | 0.0 | – | 86.0 | 40.0 | 14.2 | – | 47.7 | 6.1 | 0.7 | – |
| HbA1c tests | 36–48 | 137.4 | 63.7 | 26.0 | 9.8 | 8.3 | 1.6 | 3.5 | 0.3 | 138.8 | 48.1 | 17.7 | 9.2 | 55.9 | 4.8 | 1.7 | 0.1 |
| HbA1c tests | 60–72 | 103.5 | 60.2 | 23.8 | 9.2 | 7.2 | 0.6 | 0.0 | 0.0 | 88.3 | 34.5 | 11.8 | 8.9 | 29.4 | 4.8 | 1.0 | 0.0 |
| HbA1c tests | 84–96 | 131.1 | 95.2 | 37.8 | 10.9 | 5.1 | 1.0 | 0.0 | 0.0 | 93.4 | 48.9 | 17.6 | 9.1 | 30.8 | 4.7 | 1.7 | 0.1 |

Note: Each cell reports the squared robust first-stage $z$ statistic. Values of at least 10 are in bold. Column horizons and outcome windows are months after the index HbA1c test. All temporally eligible statistics are shown, including values below 10. Dashes denote excluded specifications in which the treatment window overlaps the outcome window. The IV figures separately apply the $z^2 \geq 10$ screen. This statistic is a one-instrument robust Wald diagnostic, not a Kleibergen–Paap F statistic, and the screen does not guarantee strong identification. Bandwidths and available samples vary across specifications. Among 144 eligible specifications per treatment definition, 115 diagnosis, 4 prescription, 106 either, and 29 both specifications have $z^2 \geq 10$ .

**Fig. A10.**
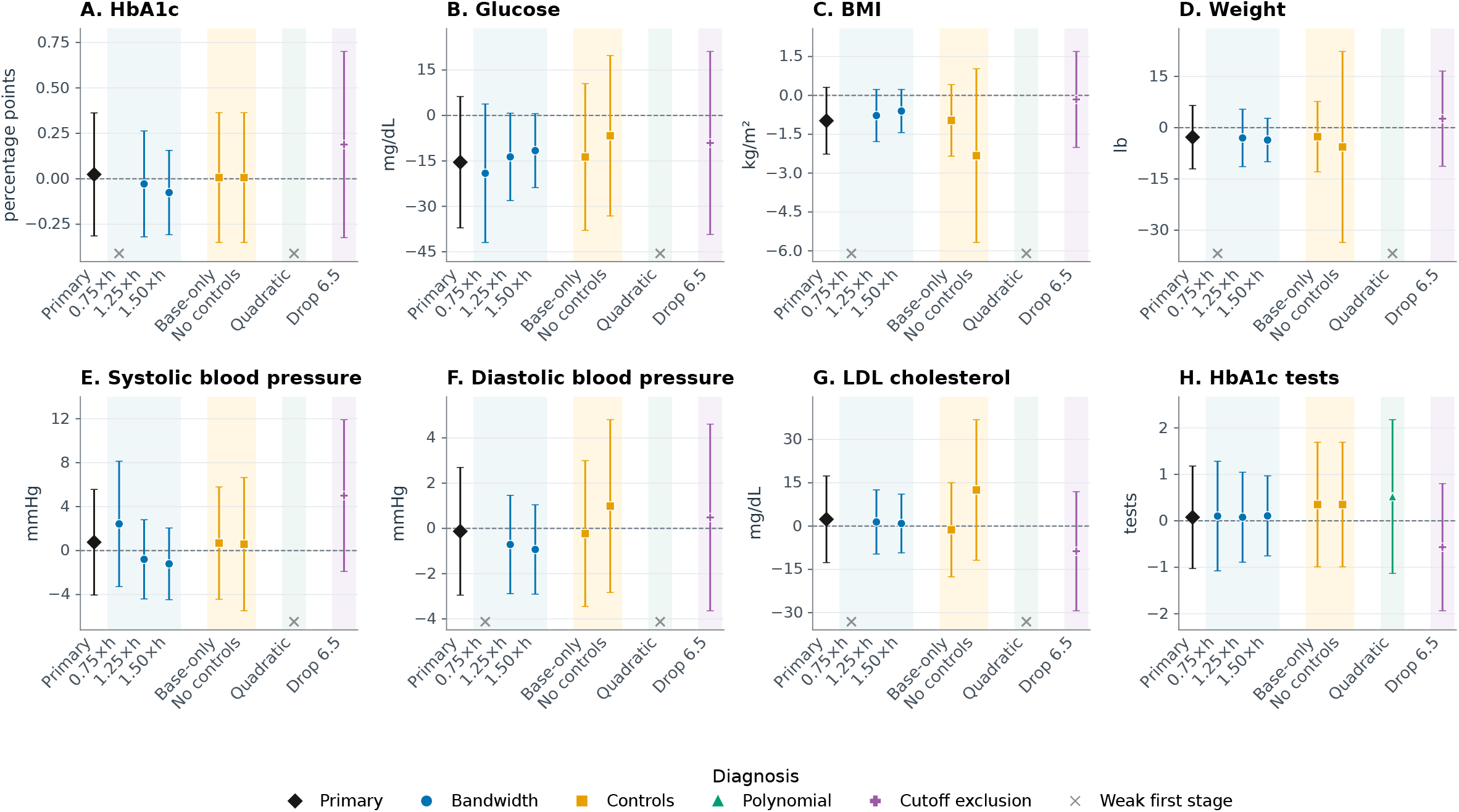
Sensitivity of primary AFC estimates: Diabetes diagnosis. Outcome averages for months 12–24 following the first HbA1c test *Note*: Panels report bias-corrected fuzzy regression-discontinuity estimates for a recorded diabetes diagnosis within six months of the index HbA1c test and outcomes measured during months 12–24. Capped vertical lines are robust 95% confidence intervals. The black diamond is the primary local-linear specification with MSE-selected bandwidth, triangular kernel weights, mass-point adjustment, controls for sex, race, ethnicity, state, index year, age, and age squared, plus the outcome’s baseline value and a baseline-missing indicator when available; standard errors are clustered by care site. Blue circles use 0.75, 1.25, and 1.50 times the primary bandwidth. Gold squares use baseline-only controls or no controls. The green triangle uses a local-quadratic specification. The purple pentagon excludes observations with index HbA1c exactly equal to 6.5%. Gray crosses along the bottom of a panel identify specifications for which the squared robust first-stage *z* statistic is below 10; their treatment estimates and intervals are suppressed. This statistic is not a Kleibergen–Paap F statistic, and the screen does not guarantee strong identification. Across the primary and seven alternative specifications for each outcome, 52 of 64 estimates pass the first-stage screen.

**Fig. A11.**
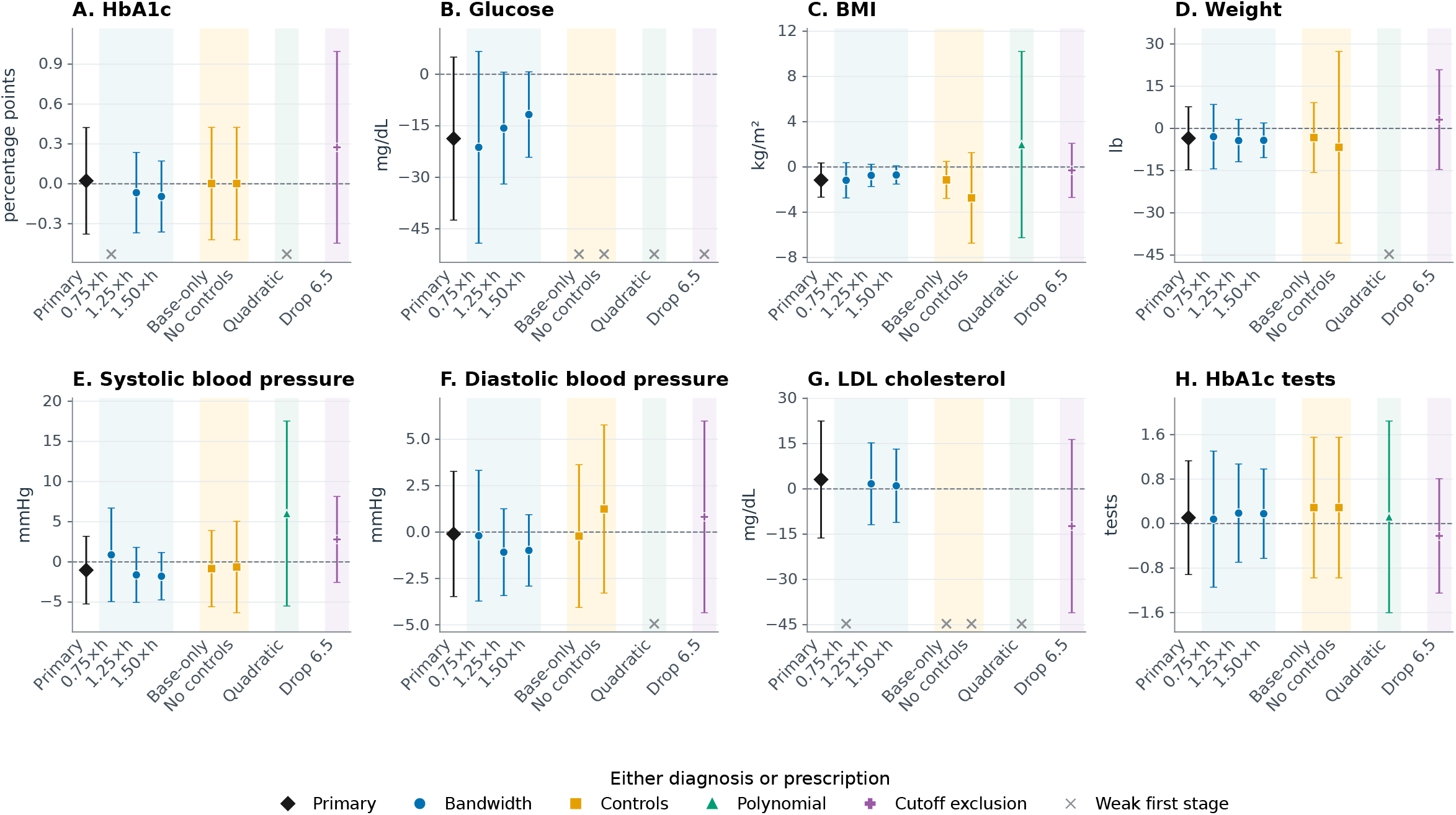
Sensitivity of primary AFC estimates: Either diabetes diagnosis or medication prescription. Outcome averages for months 12–24 following the first HbA1c test Note: Panels report bias-corrected fuzzy regression-discontinuity estimates for either a recorded diabetes diagnosis or a diabetes-medication prescription within six months of the index HbA1c test and outcomes measured during months 12–24. Capped vertical lines are robust 95% confidence intervals. The black diamond is the primary local-linear specification with MSE-selected bandwidth, triangular kernel weights, mass-point adjustment, controls for sex, race, ethnicity, state, index year, age, and age squared, plus the outcome’s baseline value and a baseline-missing indicator when available; standard errors are clustered by care site. Blue circles use 0.75, 1.25, and 1.50 times the primary bandwidth. Gold squares use baseline-only controls or no controls. The green triangle uses a local-quadratic specification. The purple pentagon excludes observations with index HbA1c exactly equal to 6.5%. Gray crosses along the bottom of a panel identify specifications for which the squared robust first-stage z statistic is below 10; their treatment estimates and intervals are suppressed. This statistic is not a Kleibergen–Paap F statistic, and the screen does not guarantee strong identification. Across the primary and seven alternative specifications for each outcome, 52 of 64 estimates pass the first-stage screen.

**Fig. A12.**
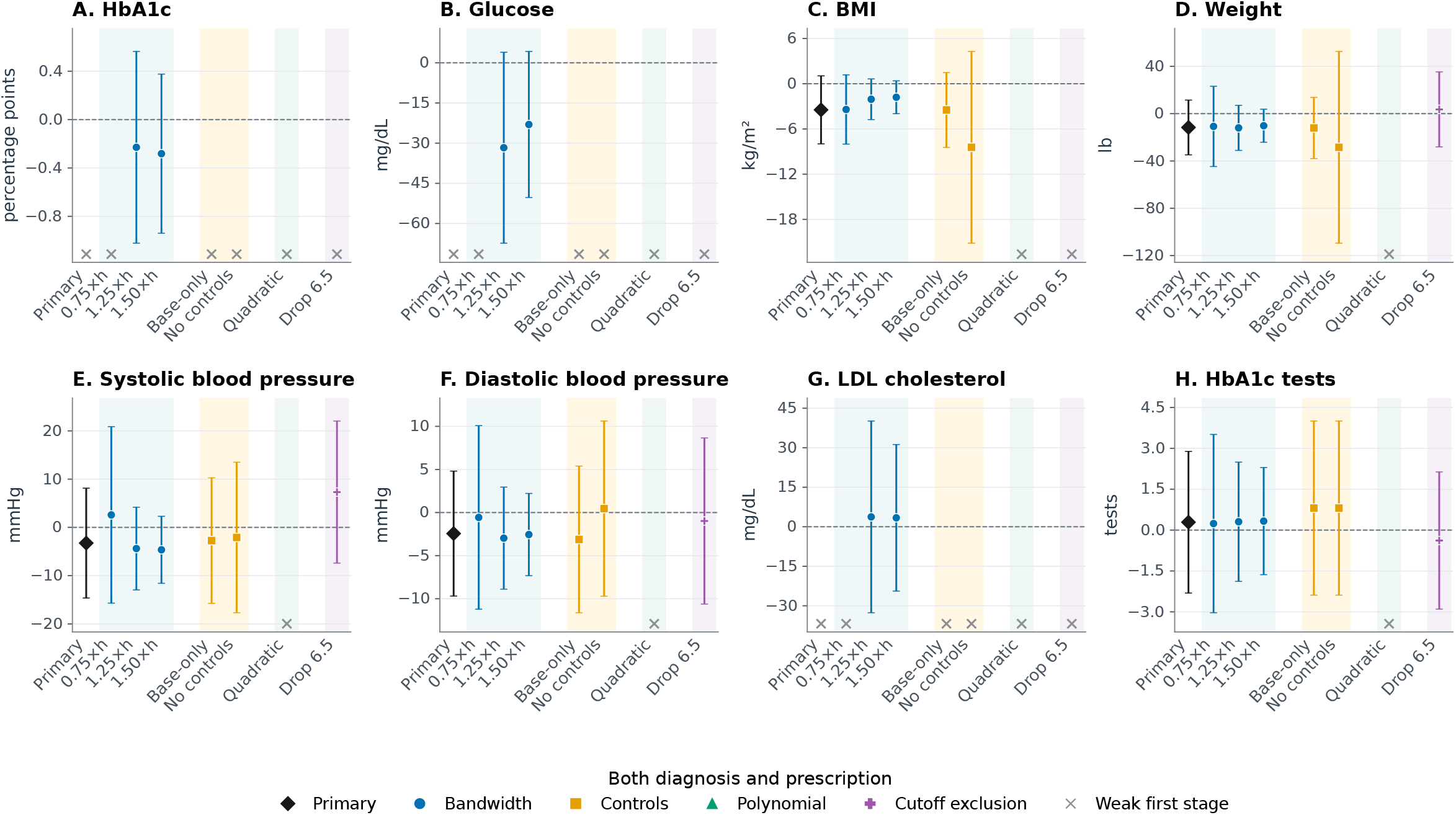
Sensitivity of primary AFC estimates: Both diabetes diagnosis and medication prescription. Outcome averages for months 12–24 following the first HbA1c test Note: Panels report bias-corrected fuzzy regression-discontinuity estimates for both a recorded diabetes diagnosis and a diabetes-medication prescription within six months of the index HbA1c test and outcomes measured during months 12–24. Capped vertical lines are robust 95% confidence intervals. The black diamond is the primary local-linear specification with MSE-selected bandwidth, triangular kernel weights, mass-point adjustment, controls for sex, race, ethnicity, state, index year, age, and age squared, plus the outcome’s baseline value and a baseline-missing indicator when available; standard errors are clustered by care site. Blue circles use 0.75, 1.25, and 1.50 times the primary bandwidth. Gold squares use baseline-only controls or no controls. The green triangle uses a local-quadratic specification. The purple pentagon excludes observations with index HbA1c exactly equal to 6.5%. Gray crosses along the bottom of a panel identify specifications for which the squared robust first-stage z statistic is below 10; their treatment estimates and intervals are suppressed. This statistic is not a Kleibergen–Paap F statistic, and the screen does not guarantee strong identification. Across the primary and seven alternative specifications for each outcome, 40 of 64 estimates pass the first-stage screen. This figure is therefore primarily a diagnostic of weak local identification rather than evidence about prescription effects.

**Fig. A13.**
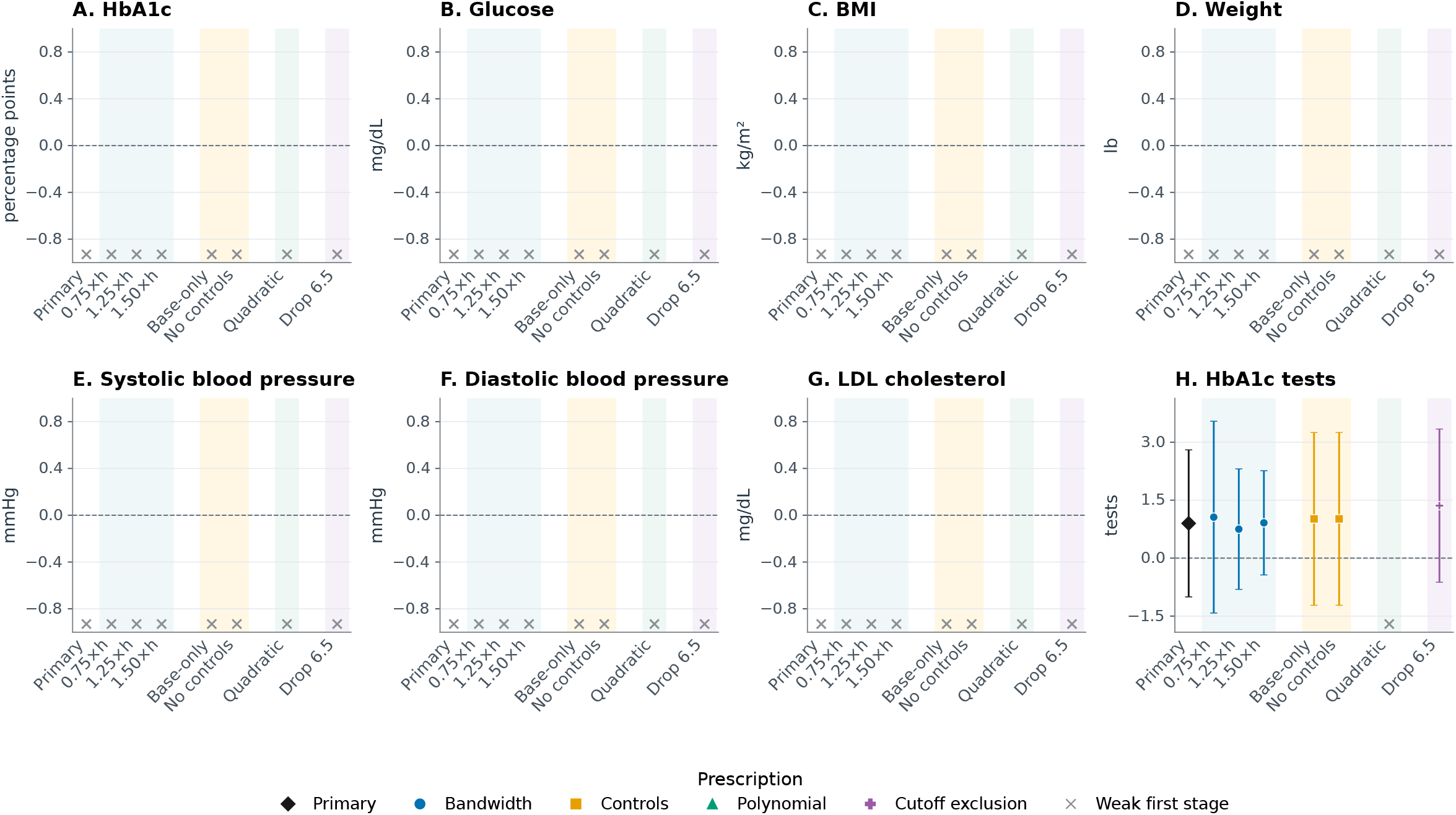
Sensitivity of primary AFC estimates: Diabetes-medication prescription. Outcome averages for months 12–24 following the first HbA1c test *Note*: Panels report bias-corrected fuzzy regression-discontinuity estimates for a diabetes-medication prescription within six months of the index HbA1c test and outcomes measured during months 12–24. Capped vertical lines are robust 95% confidence intervals. The black diamond is the primary local-linear specification with MSE-selected bandwidth, triangular kernel weights, mass-point adjustment, controls for sex, race, ethnicity, state, index year, age, and age squared, plus the outcome’s baseline value and a baseline-missing indicator when available; standard errors are clustered by care site. Blue circles use 0.75, 1.25, and 1.50 times the primary bandwidth. Gold squares use baseline-only controls or no controls. The green triangle uses a local-quadratic specification. The purple pentagon excludes observations with index HbA1c exactly equal to 6.5%. Gray crosses along the bottom of a panel identify specifications for which the squared robust first-stage *z* statistic is below 10; their treatment estimates and intervals are suppressed. This statistic is not a Kleibergen–Paap F statistic, and the screen does not guarantee strong identification. Only 7 of 64 primary and alternative estimates pass the first-stage screen. This figure is therefore primarily a diagnostic of weak local identification rather than evidence about prescription effects.

**Table A5.** American Family Cohort (AFC): Baseline continuity checks at the HbA1c diagnostic threshold.

| Outcome | Robust estimate<br>(1) | Robust 95% CI<br>(2) | Selected bandwidth<br>(3) | *N* within bandwidth<br>(4) |
| --- | --- | --- | --- | --- |
| **Panel A. Baseline outcome level** |  |  |  |  |
| HbA1c | — | — | — | — |
| Glucose | 0.498 | [-1.508, 2.504] | 0.459 | 37,745 |
| BMI | -0.397** | [-0.770, -0.023] | 0.594 | 100,953 |
| Weight | -1.463 | [-5.497, 2.571] | 0.639 | 131,446 |
| Systolic blood pressure | 0.034 | [-0.690, 0.758] | 0.585 | 97,788 |
| Diastolic blood pressure | 0.229 | [-0.251, 0.709] | 0.786 | 168,849 |
| LDL cholesterol | 0.536 | [-2.289, 3.361] | 0.702 | 117,061 |
| HbA1c tests | — | — | — | — |
| **Panel B. Baseline-outcome availability** |  |  |  |  |
| HbA1c | — | — | — | — |
| Glucose | 0.003 | [-0.075, 0.081] | 1.209 | 518,405 |
| BMI | -0.003 | [-0.040, 0.034] | 0.769 | 233,527 |
| Weight | -0.001 | [-0.047, 0.045] | 0.813 | 292,412 |
| Systolic blood pressure | -0.002 | [-0.047, 0.044] | 0.694 | 181,091 |
| Diastolic blood pressure | -0.002 | [-0.047, 0.043] | 0.697 | 181,094 |
| LDL cholesterol | -0.009 | [-0.078, 0.061] | 0.791 | 233,577 |
| HbA1c tests | — | — | — | — |

*Note:* Each row reports a bias-corrected sharp regression-discontinuity estimate at the HbA1c threshold of 6.5% with a robust 95% confidence interval. Panel A tests continuity in the observed baseline level of the outcome; Panel B tests continuity in whether the corresponding baseline measure is available. Specifications are local linear with local-quadratic bias correction, triangular kernel weights, MSE-selected bandwidths, mass-point adjustment, no covariates, and standard errors clustered by care site. *N* is the number of observations within the selected estimation bandwidth. Dashes indicate that no corresponding baseline measure was available. \* $p < 0.10$ , \*\* $p < 0.05$ , \*\*\* $p < 0.01$ , based on robust *p*-values. Intervals and significance tests are not adjusted for multiple testing.

**Table A6.**
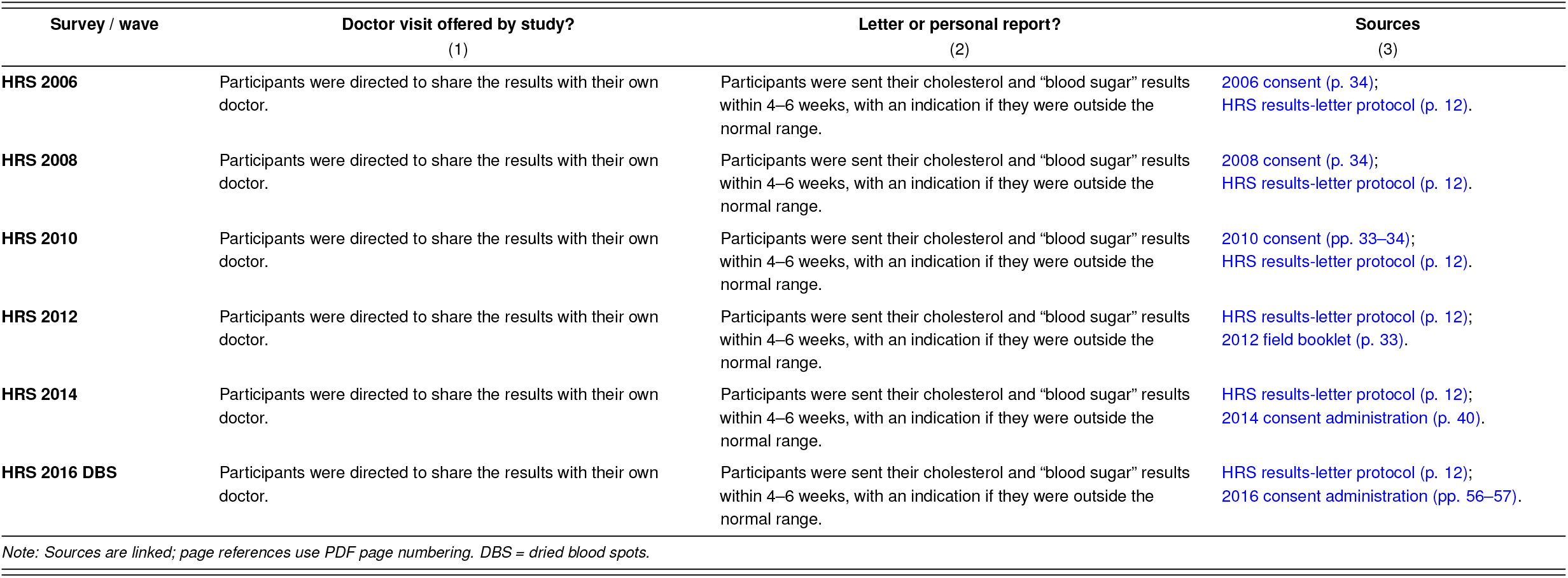
Study-provided medical visits and return of biomarker results, by survey and wave.

**Table A7.** Health and Retirement Study reduced-form results at the 6.5% cutoff.

| Outcome | 2 years |  | 4 years |  | 6 years |  | 8 years |  |
| --- | --- | --- | --- | --- | --- | --- | --- | --- |
|  | (1) |  | (2) |  | (3) |  | (4) |  |
| | $\hat{\beta}$<br>(SE) | $h$<br>$N_{\text{eff}}$ | $\hat{\beta}$<br>(SE) | $h$<br>$N_{\text{eff}}$ | $\hat{\beta}$<br>(SE) | $h$<br>$N_{\text{eff}}$ | $\hat{\beta}$<br>(SE) | $h$<br>$N_{\text{eff}}$ |
| Completely retired | 0.021<br>(0.034) | 0.883<br>3,582/316 | -0.020<br>(0.040) | 0.786<br>2,382/278 | **-0.066\***<br>(0.036) | 0.835<br>2,738/250 | -0.003<br>(0.043) | 0.830<br>2,314/225 |
| Retired in some capacity | -0.013<br>(0.038) | 0.881<br>3,582/316 | -0.005<br>(0.036) | 0.951<br>4,147/300 | **-0.076\***<br>(0.046) | 0.819<br>2,738/250 | **-0.099\***<br>(0.051) | 0.736<br>1,760/219 |
| Working | **0.077\*\***<br>(0.038) | 0.949<br>4,631/329 | -0.044<br>(0.042) | 0.651<br>1,743/264 | 0.011<br>(0.038) | 0.990<br>3,493/261 | 0.054<br>(0.054) | 0.537<br>945/201 |
| Hours worked | 1.557<br>(1.638) | 0.808<br>3,520/310 | 0.591<br>(1.503) | 0.758<br>2,322/275 | -0.432<br>(1.694) | 0.725<br>2,005/237 | 0.458<br>(1.762) | 0.817<br>2,255/218 |
| In the labor force | **0.083\*\***<br>(0.038) | 0.793<br>2,649/298 | -0.027<br>(0.044) | 0.694<br>1,710/257 | 0.008<br>(0.040) | 1.040<br>4,141/261 | 0.050<br>(0.054) | 0.551<br>911/197 |
| Not working and not retired | -0.047<br>(0.032) | 0.920<br>4,638/329 | 0.049<br>(0.033) | 0.604<br>1,749/266 | 0.028<br>(0.029) | 0.659<br>1,522/230 | 0.047<br>(0.036) | 0.785<br>1,760/219 |
| Not in the labor force & not retired | -0.039<br>(0.031) | 0.790<br>2,676/307 | 0.030<br>(0.031) | 0.677<br>1,749/266 | 0.026<br>(0.028) | 0.680<br>1,522/230 | **0.059\***<br>(0.035) | 0.754<br>1,760/219 |
| Limited by health | -0.045<br>(0.040) | 1.017<br>5,195/299 | 0.046<br>(0.057) | 0.611<br>1,568/235 | 0.023<br>(0.054) | 0.811<br>2,473/226 | 0.040<br>(0.059) | 0.729<br>1,597/193 |
| Log earnings | 0.604<br>(0.474) | 0.588<br>1,414/286 | 0.305<br>(0.440) | 0.814<br>3,215/287 | -0.016<br>(0.441) | 0.966<br>3,499/262 | 0.187<br>(0.401) | 0.812<br>2,314/225 |
| Receives SSDI benefits | -0.030<br>(0.022) | 0.519<br>1,414/286 | 0.030<br>(0.018) | 0.588<br>1,270/257 | -0.001<br>(0.017) | 0.543<br>1,095/223 | **0.054\*\*\***<br>(0.020) | 0.529<br>948/202 |
| Log SSDI benefits | -0.021<br>(0.195) | 0.712<br>2,676/307 | 0.028<br>(0.197) | 0.873<br>3,215/287 | 0.078<br>(0.233) | 0.671<br>1,522/230 | 0.063<br>(0.272) | 0.835<br>2,314/225 |
| Basic physical self-care/functioning | 0.057<br>(0.086) | 1.027<br>5,756/337 | **0.227\*\***<br>(0.097) | 0.568<br>1,269/257 | 0.151<br>(0.126) | 0.635<br>1,521/230 | 0.023<br>(0.115) | 0.970<br>2,948/236 |
| Independent everyday functioning | 0.061<br>(0.077) | 0.924<br>4,635/329 | **0.311\*\***<br>(0.121) | 0.530<br>1,269/257 | -0.072<br>(0.104) | 0.694<br>1,521/230 | -0.246<br>(0.153) | 0.501<br>946/201 |
| Self-reported health | 0.055<br>(0.097) | 1.020<br>5,753/337 | -0.085<br>(0.114) | 0.792<br>2,380/278 | 0.029<br>(0.122) | 0.625<br>1,518/229 | 0.007<br>(0.119) | 0.628<br>1,304/211 |
| Cognitive score | **-0.788\***<br>(0.443) | 0.796<br>2,607/293 | 0.545<br>(0.431) | 0.724<br>2,281/265 | -0.581<br>(0.462) | 0.878<br>2,612/238 | 0.407<br>(0.471) | 0.662<br>1,240/200 |
| Days hospitalized the past 2 years | -0.121<br>(0.094) | 0.872<br>3,550/315 | -0.001<br>(0.102) | 0.661<br>1,728/263 | 0.034<br>(0.148) | 0.794<br>2,026/237 | 0.050<br>(0.169) | 0.588<br>934/198 |
| Married | 0.006<br>(0.026) | 0.785<br>2,676/307 | -0.018<br>(0.026) | 0.741<br>2,382/278 | -0.017<br>(0.023) | 0.868<br>2,738/250 | 0.032<br>(0.038) | 0.576<br>948/202 |

*Note:* This table reports bias-corrected local-linear regression-discontinuity estimates for Health and Retirement Study respondents aged 50 or older with no prior diabetes diagnosis or diabetes medication use. Robust standard errors appear in parentheses under the coefficients. The estimates use triangular kernel weights and adjust for mass points in HbA1c. Covariates include the polygenic score for HbA1c; an indicator for missing genetic data; age at first test and its square; indicators for whether the respondent's father and mother are alive; and the natural logarithm of baseline earnings plus one, allowing zero earnings. $h$ is the bandwidth, which is the same below and above the cutoff. $N_{\text{eff}}$ gives the effective observations below/above the cutoff. The outcomes Basic physical self-care/functioning and Independent everyday functioning come from the ADL5A and IADL5A variables, respectively. ADL5A measures activities such as bathing, dressing, eating, walking across a room, and getting in and out of bed; IADL5A measures activities such as managing medication and finances and shopping. \* $p < 0.10$ , \*\* $p < 0.05$ , and \*\*\* $p < 0.01$ , based on bias-corrected $p$ -values.

**Table A8.**
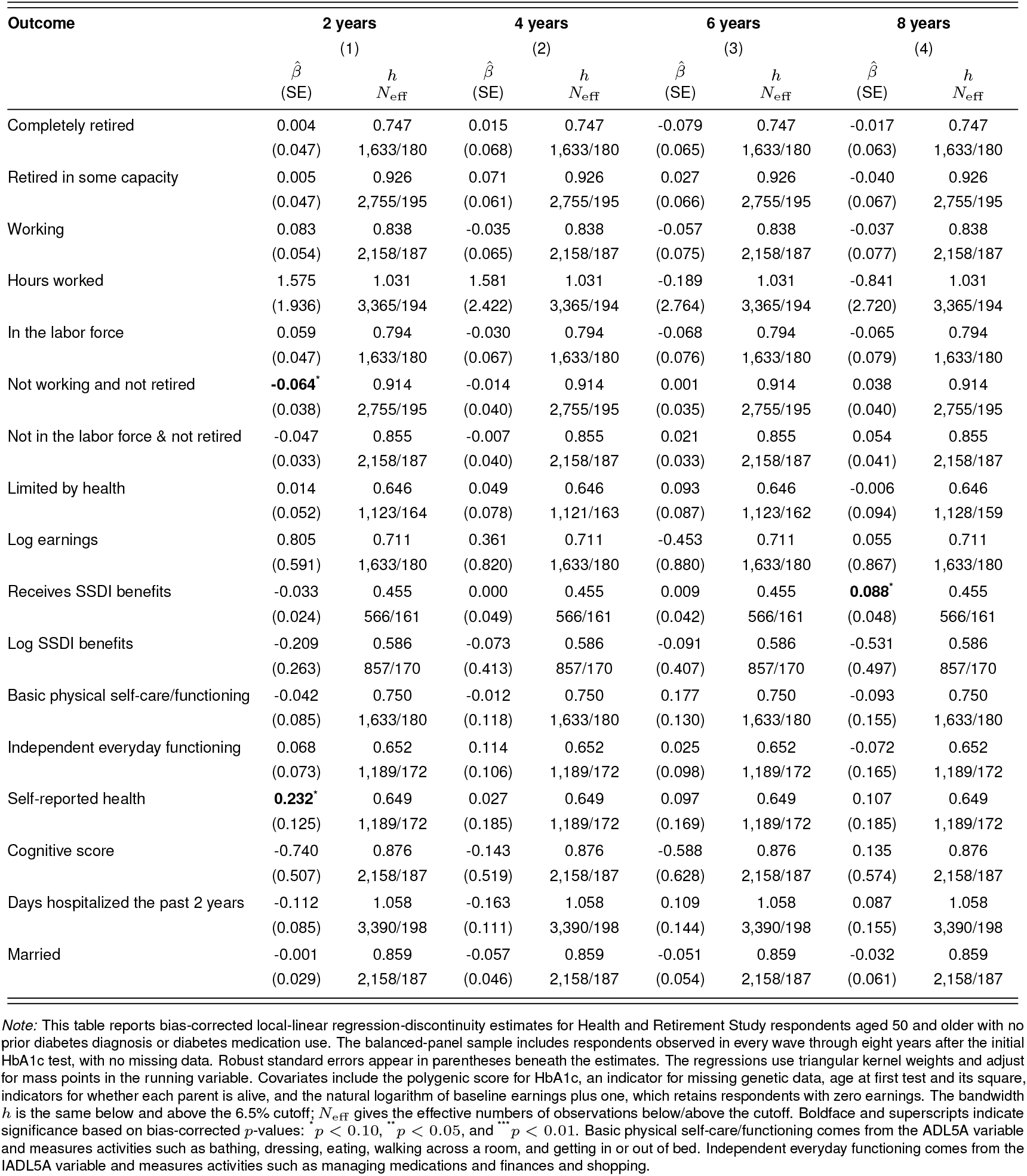
Health and Retirement Study balanced-panel reduced-form results at the 6.5% cutoff.

**Table A9.** Health and Retirement Study balanced-panel reduced-form results at the 7.0% cutoff.

| Outcome | 2 years |  | 4 years |  | 6 years |  | 8 years |  |
| --- | --- | --- | --- | --- | --- | --- | --- | --- |
|  | (1) |  | (2) |  | (3) |  | (4) |  |
| | $\hat{\beta}$<br>(SE) | $h$<br>$N_{\text{eff}}$ | $\hat{\beta}$<br>(SE) | $h$<br>$N_{\text{eff}}$ | $\hat{\beta}$<br>(SE) | $h$<br>$N_{\text{eff}}$ | $\hat{\beta}$<br>(SE) | $h$<br>$N_{\text{eff}}$ |
| Completely retired | -0.045<br>(0.066) | 1.219<br>1,794/53 | -0.023<br>(0.104) | 1.219<br>1,794/53 | -0.076<br>(0.114) | 1.219<br>1,794/53 | -0.031<br>(0.072) | 1.219<br>1,794/53 |
| Retired in some capacity | **0.217\***<br>(0.128) | 1.085<br>1,018/48 | 0.140<br>(0.154) | 1.085<br>1,018/48 | 0.008<br>(0.166) | 1.085<br>1,018/48 | -0.014<br>(0.141) | 1.085<br>1,018/48 |
| Working | **-0.365\*\*\***<br>(0.129) | 0.966<br>727/45 | -0.173<br>(0.146) | 0.966<br>727/45 | -0.013<br>(0.151) | 0.966<br>727/45 | 0.003<br>(0.136) | 0.966<br>727/45 |
| Hours worked | **-14.950\*\*\***<br>(5.085) | 0.971<br>721/45 | -4.410<br>(5.683) | 0.971<br>721/45 | 0.242<br>(5.319) | 0.971<br>721/45 | -0.816<br>(5.073) | 0.971<br>721/45 |
| In the labor force | **-0.357\*\*\***<br>(0.123) | 1.108<br>1,350/52 | -0.202<br>(0.134) | 1.108<br>1,350/52 | -0.158<br>(0.141) | 1.108<br>1,350/52 | -0.008<br>(0.132) | 1.108<br>1,350/52 |
| Not working and not retired | -0.010<br>(0.093) | 1.586<br>3,551/58 | -0.010<br>(0.094) | 1.586<br>3,551/58 | 0.072<br>(0.090) | 1.586<br>3,551/58 | 0.006<br>(0.089) | 1.586<br>3,551/58 |
| Not in the labor force & not retired | 0.043<br>(0.099) | 1.215<br>1,794/53 | -0.002<br>(0.096) | 1.215<br>1,794/53 | 0.064<br>(0.092) | 1.215<br>1,794/53 | -0.027<br>(0.099) | 1.215<br>1,794/53 |
| Limited by health | 0.095<br>(0.129) | 1.040<br>967/46 | 0.016<br>(0.139) | 1.040<br>960/47 | -0.093<br>(0.144) | 1.040<br>967/46 | -0.017<br>(0.128) | 1.040<br>965/46 |
| Log earnings | -1.555<br>(1.430) | 1.050<br>1,018/48 | -1.773<br>(1.316) | 1.050<br>1,018/48 | -1.085<br>(1.521) | 1.050<br>1,018/48 | -0.625<br>(1.499) | 1.050<br>1,018/48 |
| Receives SSDI benefits | 0.103<br>(0.089) | 1.151<br>1,350/52 | 0.072<br>(0.113) | 1.151<br>1,350/52 | 0.059<br>(0.112) | 1.151<br>1,350/52 | 0.101<br>(0.102) | 1.151<br>1,350/52 |
| Log SSDI benefits | 1.049<br>(1.091) | 1.179<br>1,350/52 | 0.187<br>(1.004) | 1.179<br>1,350/52 | -0.617<br>(0.675) | 1.179<br>1,350/52 | -0.547<br>(0.719) | 1.179<br>1,350/52 |
| Basic physical self-care/functioning | 0.052<br>(0.196) | 1.280<br>1,794/53 | -0.259<br>(0.162) | 1.280<br>1,794/53 | 0.047<br>(0.320) | 1.280<br>1,794/53 | -0.361<br>(0.278) | 1.280<br>1,794/53 |
| Independent everyday functioning | -0.058<br>(0.162) | 1.105<br>1,350/52 | -0.292<br>(0.193) | 1.105<br>1,350/52 | 0.016<br>(0.165) | 1.105<br>1,350/52 | 0.185<br>(0.348) | 1.105<br>1,350/52 |
| Self-reported health | 0.122<br>(0.181) | 1.368<br>2,318/56 | -0.198<br>(0.273) | 1.368<br>2,318/56 | 0.393<br>(0.254) | 1.368<br>2,318/56 | -0.087<br>(0.217) | 1.368<br>2,318/56 |
| Cognitive score | 0.126<br>(1.048) | 1.049<br>1,018/48 | 0.281<br>(1.288) | 1.049<br>1,018/48 | 0.144<br>(1.328) | 1.049<br>1,018/48 | -1.705<br>(1.110) | 1.049<br>1,018/48 |
| Days hospitalized the past 2 years | 0.233<br>(0.253) | 1.253<br>1,794/53 | -0.061<br>(0.191) | 1.253<br>1,794/53 | **0.557\***<br>(0.315) | 1.253<br>1,794/53 | 0.110<br>(0.288) | 1.253<br>1,794/53 |
| Married | 0.127<br>(0.090) | 1.201<br>1,794/53 | 0.173<br>(0.109) | 1.201<br>1,794/53 | 0.118<br>(0.114) | 1.201<br>1,794/53 | 0.150<br>(0.171) | 1.201<br>1,794/53 |

*Note:* This table reports bias-corrected reduced-form regression discontinuity estimates at the 7.0% HbA1c cutoff for Health and Retirement Study respondents aged 50 and older who had no prior diabetes diagnosis or diabetes medication use. The balanced-panel sample includes respondents observed in every wave through eight years after the initial HbA1c test, with no missing data. Robust standard errors appear in parentheses below the estimates. $h$ is the selected bandwidth, equal on both sides of the cutoff; $N_{\text{eff}}$ gives effective observations below/above the cutoff. Regressions use local-linear fits, triangular kernel weights, and a mass-point adjustment for the running variable. Covariates include the polygenic score for HbA1c, an indicator for missing genetic data, age at first test and its square, indicators for whether the respondent's father and mother are alive, and the natural logarithm of baseline earnings plus one, which permits respondents with zero earnings. The displayed "Basic physical self-care/functioning" outcome comes from ADL5A and measures activities such as bathing, dressing, eating, walking across a room, and getting in or out of bed. The displayed "Independent everyday functioning" outcome comes from IADL5A and measures activities such as managing medications and finances and shopping. \* $p < 0.10$ , \*\* $p < 0.05$ , and \*\*\* $p < 0.01$ , based on bias-corrected $p$ -values.

**Table A10.** Health and Retirement Study control-variable discontinuities.

| Control variable | 6.5% cutoff<br>(1) | 7.0% cutoff<br>(2) |
| --- | --- | --- |
| Polygenic score for HbA1c | -0.025<br>(0.115) | -0.104<br>(0.182) |
| Missing indicator for PGS HbA1c | **-0.135<sup>\*\*\*</sup>**<br>(0.044) | -0.167<br>(0.122) |
| Age | -0.213<br>(1.510) | 0.944<br>(2.838) |
| Age squared | -8.588<br>(216.488) | 118.644<br>(415.477) |
| Whether respondent's father is alive | -0.020<br>(0.048) | 0.096<br>(0.083) |
| Whether respondent's mother is alive | -0.021<br>(0.056) | **0.251<sup>\*\*</sup>**<br>(0.109) |
| Baseline log earnings | -0.788<br>(0.763) | -0.673<br>(1.128) |

*Note:* This table reports bias-corrected regression discontinuity estimates for predetermined control variables among Health and Retirement Study respondents aged 50 and older who had no prior diabetes diagnosis or diabetes medication use. Robust standard errors appear in parentheses. The regressions use triangular kernel weights, adjust for mass points in the running variable, and include no covariates. Results use the two-year follow-up. \* $p < 0.10$ , \*\* $p < 0.05$ , and \*\*\* $p < 0.01$ , based on bias-corrected $p$ -values.

**Table A11.** Health and Retirement Study first-stage results.

| **First-stage outcome** | **6.5% cutoff**<br>**(1)** | **7.0% cutoff**<br>**(2)** |
| --- | --- | --- |
| Self-reported diabetes diagnosis | 0.021<br>(0.036) | -0.037<br>(0.096) |
| Self-reported diabetes medication use | -0.021<br>(0.027) | 0.076<br>(0.082) |

*Note:* This table reports bias-corrected first-stage regression discontinuity estimates for Health and Retirement Study respondents aged 50 and older who had no prior diabetes diagnosis or diabetes medication use. Robust standard errors appear in parentheses. The regressions use triangular kernel weights and adjust for mass points in the running variable. Covariates include the polygenic score for HbA1c, an indicator for missing genetic data, age at first test and its square, indicators for whether the respondent's father and mother are alive, and the natural logarithm of baseline earnings plus one, which permits the inclusion of respondents with zero earnings. Results use the two-year follow-up. Diagnosis and medication use are self-reported. \* $p < 0.10$ , \*\* $p < 0.05$ , and \*\*\* $p < 0.01$ , based on bias-corrected $p$ -values.

**Table A12.** Health and Retirement Study robustness checks at the 6.5% cutoff.

| Outcome | Baseline placebo<br>(1) | Baseline outcome only<br>(2) | No covariates<br>(3) | Exclude cutoff<br>(4) | Cap at 6.9%<br>(5) | 0.75× bandwidth<br>(6) | 1.25× bandwidth<br>(7) |
| --- | --- | --- | --- | --- | --- | --- | --- |
| Completely retired | -0.016<br>(0.040) | 0.015<br>(0.033) | 0.003<br>(0.038) | 0.032<br>(0.054) | 0.023<br>(0.039) | 0.017<br>(0.050) | 0.021<br>(0.036) |
| Retired in some capacity | -0.035<br>(0.049) | -0.025<br>(0.042) | -0.040<br>(0.061) | -0.004<br>(0.067) | -0.005<br>(0.044) | 0.052<br>(0.055) | -0.009<br>(0.041) |
| Working | -0.033<br>(0.039) | **0.082<sup>\*\*</sup>**<br>(0.037) | 0.042<br>(0.064) | 0.081<br>(0.060) | **0.085<sup>\*</sup>**<br>(0.048) | 0.056<br>(0.054) | **0.077<sup>\*</sup>**<br>(0.041) |
| Hours worked | 0.335<br>(1.894) | 1.574<br>(1.743) | -1.067<br>(2.930) | 2.714<br>(2.800) | 0.794<br>(1.943) | -1.537<br>(2.444) | 1.195<br>(1.750) |
| In the labor force | -0.009<br>(0.041) | **0.083<sup>\*\*</sup>**<br>(0.037) | 0.068<br>(0.064) | 0.087<br>(0.067) | **0.084<sup>\*</sup>**<br>(0.045) | 0.002<br>(0.060) | **0.083<sup>\*\*</sup>**<br>(0.041) |
| Not working and not retired | 0.035<br>(0.042) | -0.034<br>(0.032) | -0.029<br>(0.034) | -0.014<br>(0.058) | **-0.069<sup>\*</sup>**<br>(0.037) | **-0.085<sup>\*</sup>**<br>(0.049) | -0.049<br>(0.034) |
| Not in the labor force and not retired | -0.003<br>(0.037) | -0.023<br>(0.031) | -0.035<br>(0.031) | 0.009<br>(0.056) | **-0.078<sup>\*\*</sup>**<br>(0.035) | -0.064<br>(0.052) | **-0.056<sup>\*</sup>**<br>(0.034) |
| Limited by health | -0.007<br>(0.053) | -0.048<br>(0.044) | 0.007<br>(0.058) | **-0.147<sup>\*\*</sup>**<br>(0.063) | -0.008<br>(0.050) | -0.013<br>(0.057) | -0.041<br>(0.043) |
| Log earnings | -0.000<br>(0.000) | 0.669<br>(0.470) | 0.236<br>(0.577) | 0.735<br>(0.793) | 0.115<br>(0.609) | -0.201<br>(0.822) | 0.426<br>(0.535) |
| Receives Social Security Disability benefits | -0.014<br>(0.022) | -0.033<br>(0.023) | -0.038<br>(0.031) | -0.053<br>(0.047) | -0.042<br>(0.027) | -0.080<br>(0.054) | -0.038<br>(0.026) |
| Log Social Security Disability benefits | -0.223<br>(0.253) | 0.040<br>(0.200) | -0.031<br>(0.263) | 0.097<br>(0.348) | -0.040<br>(0.200) | 0.015<br>(0.336) | 0.027<br>(0.222) |
| ADL5A (basic physical self-care/functioning) | 0.005<br>(0.100) | 0.067<br>(0.087) | 0.046<br>(0.127) | -0.055<br>(0.152) | 0.041<br>(0.103) | 0.014<br>(0.122) | 0.044<br>(0.093) |
| IADL5A (independent everyday functioning) | -0.071<br>(0.089) | 0.083<br>(0.083) | 0.116<br>(0.113) | 0.018<br>(0.135) | 0.054<br>(0.089) | 0.092<br>(0.107) | 0.041<br>(0.081) |
| Self-reported health | 0.154<br>(0.127) | 0.060<br>(0.098) | 0.182<br>(0.119) | -0.061<br>(0.169) | 0.108<br>(0.116) | 0.109<br>(0.138) | 0.064<br>(0.103) |
| Cognitive score | 0.162<br>(0.471) | **-0.789<sup>\*</sup>**<br>(0.441) | -0.481<br>(0.518) | -1.049<br>(0.730) | -0.790<br>(0.505) | **-1.440<sup>\*\*</sup>**<br>(0.715) | **-0.825<sup>\*</sup>**<br>(0.481) |
| Days hospitalized during the past two years | **-0.213<sup>\*\*</sup>**<br>(0.083) | -0.135<br>(0.104) | **-0.261<sup>\*\*</sup>**<br>(0.114) | 0.004<br>(0.158) | -0.156<br>(0.104) | -0.020<br>(0.158) | -0.140<br>(0.103) |
| Married | -0.078<br>(0.055) | 0.006<br>(0.026) | -0.073<br>(0.057) | -0.004<br>(0.035) | 0.015<br>(0.033) | 0.039<br>(0.041) | 0.009<br>(0.028) |

Note: Each cell reports a bias-corrected reduced-form regression-discontinuity estimate, with the robust standard error in parentheses on the next row. Unless a column specifies otherwise, regressions use triangular kernel weights, adjust for mass points in the running variable, and include the polygenic score for HbA1c, an indicator for missing genetic data, age at first test and its square, indicators for whether the respondent's father and mother are alive, and the natural logarithm of baseline earnings plus one. Results use the outcome two years after the initial HbA1c test, except in column (1). Column (1), Baseline placebo, uses the baseline value of each outcome instead and retains the usual controls. Column (2), Baseline outcome only, keeps the two-year outcome and uses its baseline value as the sole control. Column (3), No covariates, omits all controls, including the baseline outcome. Column (4), Exclude cutoff, drops observations with HbA1c exactly equal to 6.5%. Column (5), Cap at 6.9%, excludes observations at or above 7.0%. Columns (6) and (7) multiply the optimal bandwidth selected by Stata's `rdrobust` command by 0.75 and 1.25, respectively. <sup>\*</sup> $p < 0.10$ , <sup>\*\*</sup> $p < 0.05$ , and <sup>\*\*\*</sup> $p < 0.01$ , based on bias-corrected $p$ -values. Confidence intervals, sample counts, effective sample counts, and bandwidths are retained in the accompanying Excel workbooks.

**Table A13.** Health and Retirement Study robustness checks at the 7.0% cutoff.

| Outcome | Baseline placebo<br>(1) | Baseline outcome only<br>(2) | No covariates<br>(3) | Exclude cutoff<br>(4) | 0.75× bandwidth<br>(5) | 1.25× bandwidth<br>(6) |
| --- | --- | --- | --- | --- | --- | --- |
| Completely retired | -0.022<br>(0.041) | **-0.099\***<br>(0.056) | -0.094<br>(0.059) | 0.046<br>(0.134) | -0.117<br>(0.086) | **-0.118\*\***<br>(0.058) |
| Retired in some capacity | 0.026<br>(0.084) | 0.086<br>(0.083) | 0.125<br>(0.110) | 0.045<br>(0.102) | 0.099<br>(0.090) | 0.089<br>(0.072) |
| Working | 0.036<br>(0.076) | **-0.208\*\*\***<br>(0.076) | **-0.238\*\***<br>(0.116) | -0.103<br>(0.096) | -0.160<br>(0.103) | **-0.223\*\*\***<br>(0.079) |
| Hours worked | 5.038<br>(4.987) | **-6.790\***<br>(3.492) | -4.811<br>(6.657) | **-7.860\*\***<br>(3.574) | -5.821<br>(5.441) | **-6.470\***<br>(3.796) |
| In the labor force | -0.003<br>(0.071) | **-0.199\*\*\***<br>(0.074) | **-0.292\*\***<br>(0.114) | **-0.132\***<br>(0.080) | **-0.164\***<br>(0.093) | **-0.206\*\*\***<br>(0.075) |
| Not working and not retired | -0.016<br>(0.072) | 0.003<br>(0.065) | 0.016<br>(0.073) | -0.036<br>(0.080) | -0.051<br>(0.091) | -0.001<br>(0.070) |
| Not in the labor force and not retired | 0.074<br>(0.081) | 0.020<br>(0.062) | 0.050<br>(0.068) | -0.003<br>(0.074) | 0.006<br>(0.076) | 0.017<br>(0.065) |
| Limited by health | 0.040<br>(0.090) | 0.082<br>(0.080) | 0.108<br>(0.106) | 0.170<br>(0.149) | -0.067<br>(0.117) | 0.036<br>(0.087) |
| Log earnings | -0.000<br>(0.000) | -0.395<br>(0.894) | -0.764<br>(1.161) | -0.549<br>(1.210) | -0.712<br>(1.277) | -0.484<br>(0.929) |
| Receives Social Security Disability benefits | **0.119\*\***<br>(0.055) | 0.017<br>(0.050) | **0.115\***<br>(0.070) | 0.014<br>(0.067) | 0.016<br>(0.078) | 0.022<br>(0.054) |
| Log Social Security Disability benefits | **0.936\*\***<br>(0.474) | 0.214<br>(0.652) | 0.944<br>(0.665) | 0.279<br>(1.131) | 0.225<br>(0.761) | 0.132<br>(0.673) |
| ADL5A (basic physical self-care/functioning) | -0.108<br>(0.155) | -0.008<br>(0.186) | -0.151<br>(0.246) | -0.172<br>(0.222) | 0.071<br>(0.310) | -0.045<br>(0.235) |
| IADL5A (independent everyday functioning) | -0.158<br>(0.164) | -0.173<br>(0.165) | -0.296<br>(0.215) | -0.402<br>(0.325) | -0.193<br>(0.278) | -0.160<br>(0.186) |
| Self-reported health | **-0.459\***<br>(0.251) | 0.165<br>(0.172) | -0.122<br>(0.196) | 0.443<br>(0.325) | 0.151<br>(0.251) | 0.178<br>(0.178) |
| Cognitive score | 1.102<br>(0.858) | 0.233<br>(0.755) | 0.836<br>(1.041) | 0.209<br>(1.128) | -1.035<br>(0.958) | 0.221<br>(0.736) |
| Days hospitalized during the past two years | -0.096<br>(0.230) | 0.082<br>(0.208) | 0.001<br>(0.199) | -0.029<br>(0.310) | 0.165<br>(0.295) | 0.113<br>(0.218) |
| Married | 0.137<br>(0.114) | 0.045<br>(0.058) | 0.180<br>(0.116) | 0.080<br>(0.093) | 0.067<br>(0.088) | 0.054<br>(0.060) |

Note: Each cell reports a bias-corrected reduced-form regression-discontinuity estimate, with the robust standard error in parentheses on the next row. Unless a column specifies otherwise, regressions use triangular kernel weights, adjust for mass points in the running variable, and include the polygenic score for HbA1c, an indicator for missing genetic data, age at first test and its square, indicators for whether the respondent's father and mother are alive, and the natural logarithm of baseline earnings plus one. Results use the outcome two years after the initial HbA1c test, except in column (1). Column (1), Baseline placebo, uses the baseline value of each outcome instead and retains the usual controls. Column (2), Baseline outcome only, keeps the two-year outcome and uses its baseline value as the sole control. Column (3), No covariates, omits all controls, including the baseline outcome. Column (4), Exclude cutoff, drops observations with HbA1c exactly equal to 7.0%. Columns (5) and (6) multiply the optimal bandwidth selected by Stata's `rdrobust` command by 0.75 and 1.25, respectively. \* $p < 0.10$ , \*\* $p < 0.05$ , and \*\*\* $p < 0.01$ , based on bias-corrected $p$ -values. Confidence intervals, sample counts, effective sample counts, and bandwidths are retained in the accompanying Excel workbooks.

## Footnotes

* Version details: AFC OMOP RIF v5.0, DOI 10.57761/q0zb-7s60.

† The complete letter can be viewed here.

‡ A detailed prescription drug study took place in 2005 and 2007, providing more detail on prescription drug use, but for only around 25 percent of respondents.

§ In the case of diabetes, this is if the HbA1c is 6.5 or higher.

¶ Technically the squared robust first-stage z statistic.

